# From wastewater to GIS-based reporting: the ANNA-WES data model for reliable biomarker tracking in wastewater and environmental surveillance

**DOI:** 10.1101/2025.03.18.25324175

**Authors:** Anna Uchaikina, Anna-Sonia Kau, Alexander Graf, Christine Walzik, Alexander Mitranescu, Lisa Falk, Mohammad Shehryaar Khan, Claudia Stange, Johannes Ho, Katalyn Roßmann, Ingo Michels, Nathan Obermaier, Cristina J. Saravia, Susanne Rost, Thorsten Portain, Jürgen Demeter, Christopher Becker, Martina Füchsle, Fabienne Kaymaz-Ried, Alexander Klaus, Tobias Ziegler, Katharina Springer, Melissa Hohl, Peter-Louis Plaumann, Annemarie Bschorer, Stefanie Huber, Patrick Dudler, Andreas Tiehm, Jörg E. Drewes, Christian Wurzbacher

## Abstract

Since the COVID-19 pandemic, wastewater-based epidemiology (WBE) has emerged as a useful additional diagnostic tool for public health management. For rapid reporting of results and an automated implementation of WBE as a monitoring tool, a digital work flow of data is crucial. Here, we present the Automated Network for Normalization, Analysis, and Visualization of Wastewater and Environmental Surveillance (ANNA-WES) – a comprehensive workflow integrating GIS-based data entry, Python-driven data processing, and ArcGIS-supported visualization. ANNA-WES streamlines data transfer between wastewater treatment plant operators, decision-makers, and the public while ensuring harmonized data processing for transferability, precise georeferencing of index cases, and near real-time SARS-CoV-2 biomarker reporting. To enhance data reliability, we embedded an unsupervised quality control algorithm that filters outliers based on gene ratios, surrogate viruses, water quality parameters, and theoretical reproductive value thresholds. Designed for scalability, ANNA-WES integrates into public dashboards and can be combined with regional or national health data, providing a robust decision-support system for infectious disease surveillance. The workflow is adaptable to various pathogens or biomarkers, advancing WBE as a continuous, quality-controlled public health monitoring tool.

**Highlights:**

- Digitized wastewater-based epidemiology system for automated and immediate display of results
- Automated quality control algorithm based on metadata and epidemiological considerations
- Intersection of SARS-CoV-2 public health data with wastewater service areas

## 1. Introduction

Emerging in late 2019, the SARS-CoV-2 virus spread across the globe, challenging health systems worldwide with approximately 772 million infections and 7 million recorded deaths as of October 2024 (WHO, 2024). To prevent health system collapse, effective tracking of SARS-CoV-2 cases and early detection of new infection surges became essential for public health management. Since wastewater-based epidemiology (WBE) had already proven effective as an early warning system for poliovirus (Cowger et al., 2017; Kopel et al., 2014; Paul et al., 1940; WHO, 2003), different research groups considered whether SARS-CoV-2 can be monitored in wastewater. Early trials during the emergence of the COVID-19 pandemic demonstrated the successful detection and quantification of SARS-CoV-2 RNA sequences in wastewater across several countries, including the Netherlands (Lodder & Roda Husman, 2020; Medema et al., 2020a), Australia (Ahmed et al., 2020), Italy (La Rosa et al., 2021), Spain (Chavarria-Miró et al., 2021; Llanos et al., 2022), Germany (Ho et al., 2021), and the USA (Sherchan et al., 2020). Recognized as a valuable diagnostic tool, WBE provided critical complementary information for public health decision-making by revealing infection trends within sewer service areas, independent of clinical testing (EU, 2021). It was deployed on both small (Karthikeyan et al., 2021; Llanos et al., 2022; Roßmann et al., 2022; Deng et al., 2022) and large scales (Amman et al., 2022; Daleiden et al., 2022; Jmii et al., 2021; Medema et al., 2020b; Pillay et al., 2021; Keshaviah et al., 2021).

During the COVID-19 pandemic, effective sharing of SARS-CoV-2 infection data became crucial to foster stakeholder engagement and increase public acceptance of lockdown and containment measures (Naughton et al., 2023b). To enhance accessibility and improve public health communication, user-friendly dashboards were developed as effective tools for sharing data (Kamel Boulos & Geraghty, 2020). Initially, these dashboards focused on clinical COVID-19 data at global (JHU, 2023; WHO, 2024) or national levels (e. g., France (Santé Publique France, 2023), Germany (RKI, 2023b)), while WBE data was often shared only via web-based platforms (CDC, 2024; New York State, 2025; RIVM, 2024; UKHSA, 2022). Over time, dashboards evolved to incorporate SARS-CoV-2 wastewater surveillance data from regional initiatives (San Diego (SEARCH, 2025), New York (Hill et al., 2023), county of Berchtesgadener Land (Landratsamt Berchtesgadener Land, 2022), county of Augsburg (Presse Augsburg, 2023)), as well as from national efforts (South Africa (SAMCR, 2023), New Zealand (ESR, 2024), Belgium (Sciensano, 2023), Sweden (SciLifeLab & Pathogens Sweden, 2025), United States (Adams et al., 2024; CDC, 2024)). While some of these dashboards are no longer active, platforms like COVIDPoops (UC, 2025) and Wastewater Sphere (Global Water Pathogens Project, 2025) provide comprehensive overviews of global wastewater monitoring efforts. However, many WBE sampling locations, numbering in the hundreds, continued to collect wastewater data without integrating it into online dashboards (Naughton et al., 2023a).

Despite the usefulness of these dashboards in sharing data effectively, the rapid development of region-specific solutions created new challenges. Early frameworks for processing and presenting WBE data were introduced during the COVID-19 pandemic (Gonzalez et al., 2020; Mitranescu et al., 2022; Rauch et al., 2022), as local health authorities utilized SARS-CoV-2 WBE research findings to evaluate pandemic dynamics. Region-specific solutions had to be quickly developed to alleviate the manual workload for crisis response teams, who were often overwhelmed by the need to produce weekly reports (Roßmann et al., 2022).

However, the lack of harmonization across these systems increased the risk of misinterpretation, especially among stakeholders less familiar with the data structures (Naughton et al., 2023a). Larger health authorities at the state and national level faced additional challenges in comparing and assessing cross-regional data due to discrepancies in units and scales for biomarker concentration, data presentation (e.g., bar charts, line charts, tables, maps), and types of data included (e.g., clinical case data, hospitalization and mortality rates, variant monitoring) (Keshaviah et al., 2021; Naughton et al., 2023a; Ciannella et al., 2023; Adams et al., 2024). Furthermore, having data scattered across reports, local web pages, and unpublished dashboards makes valuable surveillance information harder to find (MacDonald et al., 2017), underscoring the need for globally harmonized data sharing methods and workflows in WBE processes.

To improve the practical application of WBE, data models should incorporate harmonized strategies for data entry, automated processing, cloud-based storage, and easy-to-read visuals (Kirby et al., 2021; Mendoza et al., 2023; Morgan, 2019). Real-time reporting and feedback loops are also essential, ensuring that WBE results can support rapid public health responses during a pandemic (Dong et al., 2020; Morgan, 2019). Additionally, aligning sewer service areas with boundaries of political jurisdictions is essential for accurate, population-specific comparisons, particularly in regions with decentralized systems (like septic tanks) where sewer systems do not cover the whole city population (Street et al., 2020) or when sampling represents only specific city districts. Discrepancies between sewer service areas and political boundaries can lead to inaccuracies in correlating wastewater data with clinical case numbers, which are typically reported by political jurisdictions (Castiglioni et al., 2013; Lai et al., 2011; Thomas et al., 2017; Tscharke et al., 2019).

Another challenge in wastewater surveillance is the significant variance observed in the results themselves due to their complex nature. Key contributing factors include (a) individual differences in virus shedding rates influenced by infection status and virus variants, (b) population dynamics, (c) sewer system impacts due to different discharges within the service area, (d) travel time and degradation processes in the sewer, (e) dilution effects from precipitation or non-municipal wastewater, (f) variability introduced during sampling and laboratory protocols, and (g) data evaluation methods. Together, these factors can result in variations in biomarker concentrations, challenging reliable infection trend interpretation.

Many research groups have investigated individual factors contributing to these fluctuations, focusing on optimizing and standardizing key components essential for WBE, such as laboratory procedures for RNA extraction and quantification (Ciannella et al., 2023; Oyervides-Muñoz et al., 2024; Pecson et al., 2021), statistical correlation between viral concentration and clinical testing data (Atique et al., 2021; Hillary et al., 2021; Salvagno et al., 2022), impacts of different SARS-CoV-2 variants (Agrawal et al., 2022; Brunner et al., 2022; van Vo et al., 2022), the stability of various markers for WBE (Gao et al., 2022), and sampling strategies (Augusto et al., 2022; Uchaikina et al., 2023; Wilson et al., 2022). Some WBE workflows incorporate normalization parameters, smoothing algorithms, and machine-learning approaches to mitigate these data variations. While smoothing algorithms reduce fluctuations mathematically, normalization parameters aim to link variations to specific causative factors within the sewer system. However, no single normalization factor currently accounts for and resolves all variances inherent in wastewater systems (Mitranescu et al., 2022; O’Brien et al., 2023; Saravia et al., 2024). As a result, SARS-CoV-2 WBE results are typically processed using a combination of normalization and smoothing to mitigate data variation.

Modeling approaches in WBE have demonstrated potential for predicting clinical indicators but often face limitations due to high data variability. For example, Ai et al. (2022), Galani et al. (2022), and Schenk et al. (2023) achieved promising predictive accuracy but did not address sewer network processes, which significantly impact wastewater data reliability. While these studies implicitly aim to enhance data robustness through statistical adjustments or model optimization, their quality control (QC) measures remain limited to addressing specific variables rather than the multifactorial dynamics of WBE systems. Schmid et al. (2024) developed a theoretical model incorporating sewer dynamics such as precipitation, sampling protocols, and viral decay, offering valuable insights into WBE variability. However, the lack of real-world validation and integration with comprehensive QC protocols limits its practical applicability.

Despite some progress made through programs established during the COVID-19 pandemic, a fully integrated, practical pipeline that consolidates these insights remains largely unestablished. To our knowledge, no existing approach offers a holistic, end-to-end workflow encompassing sampling, automated data processing, robust QC, and seamless reporting based on practical, real-world data. Critically, existing approaches lack automated QC protocols capable of addressing multifactorial influences across the WBE process, ensuring data reliability, and enabling actionable insights for public health decision-making (Ahmed & Bivins et al., 2020; Ciannella et al., 2023; Naughton et al., 2023a; Rauch et al., 2022). This gap underscores the need for a unified framework that integrates data processing, QC, and reporting to enhance the effectiveness of pandemic management and future surveillance efforts.

Our project bridges these gaps by developing (i) a near real-time reporting system for biomarker results to health authorities and the public, (ii) an unsupervised automatic QC algorithm to process biomarker results and filter outliers, (iii) a method to intersect public health data with wastewater service area boundaries to display only geo-matched parameters (georeferenced index cases), and (iv) a comprehensive, transferable data model grounded in our well-established workflow.

In this work, we present our workflow and data model ANNA-WES (Automated Network for Normalization, Analysis, and Visualization of Wastewater and Environmental Surveillance), and the automated QC algorithm SSQN (SARS-CoV-2 sewage quality control and normalization). Both were developed using SARS-CoV-2 biomarker data from a wastewater surveillance program conducted at eleven full-scale wastewater treatment plants (WWTPs) of varying capacities and three sewer sampling locations in Southern Germany. We also evaluate the alignment of clinical data with wastewater data based on corresponding wastewater service areas. The workflow was initially developed in collaboration with public health authorities in the county of Berchtesgadener Land, the county of Augsburg, and the Joint Medical Service of the German Armed Forces (Zentraler Sanitätsdienst der Bundeswehr) (Roßmann et al., 2021; Roßmann et al., 2022). Given the crisis situation, the workflow was initially deployed to meet immediate needs, with additional refinements following a thorough assessment based on real-time feedback. As experience accumulated, we expanded the workflow to further communities to support public health agencies in managing the COVID-19 pandemic, incorporating features such as georeferenced alignment of clinical and wastewater service area data and an automated QC algorithm – enhancements that can now be optionally integrated.

## 2. Materials and Methods

### Sampling sites, laboratory methods, and overview on data processing steps

We collected samples from eleven WWTPs and three sewer sampling sites, covering 13 communities and one city district in Southern Germany, as listed in Table 1. Sampling was conducted twice per week, with sampling campaigns spanning up to 4.5 years. A detailed overview is provided in Supplementary Information (Supplement S1), including the sample type, duration, and the data used to test the SSQN algorithm for QC analysis.

**Table 1:** List of sampling locations, population served by the WWTP, associated laboratories, and sampling periods covered by this publication (note: sampling campaigns at most locations continued after the periods covered in this study)

| Sampled community | Sampling site | Population served | Laboratory | Sampling period |
| --- | --- | --- | --- | --- |
| Munich | WWTP Gut Großlappen | 1,156,000 | TUM | 07/2021 - 12/2024 |
| Nuremberg | WWTP Nuremberg Klärwerk 1 | 589,000* | LGL (Erlangen) | 12/2022 - 12/2024 |
| Karlsruhe | WWTP Karlsruhe | 370,000 | TZW | 07/2020 - 12/2024 |
| Augsburg | WWTP Augsburg | 343,000 | TUM | 07/2021 - 12/2024 |
| Augsburg | WWTP Augsburg | 343,000 | Augsburg WWTP | 02/2022 - 07/2022 |
| Weiden | WWTP Weiden | 43,400 | TUM | 07/2021 - 12/2024 |
| Königsbrunn (Augsburg county) | Sewer manhole | 29,500 | TUM | 07/2021 - 12/2024 |
| Berchtesgaden | WWTP Berchtesgaden | 27,000 | TZW, from 03/2023 LGL (Erlangen) | 11/2020 - 12/2024 |
| Munich district | Pump station | 25,000 | TUM | 07/2021 - 05/2023 |
| Freilassing | WWTP Freilassing | 20,000 | TZW, from 03/2023 LGL (Erlangen) | 11/2020 - 12/2024 |
| Bad Reichenhall | WWTP Piding | 19,500 | TZW, from 03/2023 LGL (Erlangen) | 11/2020 - 12/2024 |
| Stadtbergen (Augsburg county) | Sewer manhole | 15,000 | TUM | 07/2021 - 12/2024 |
| Ebersberg | WWTP Ebersberg | 12,500 | LGL (Oberschleißheim) | 04/2023 - 12/2024 |
| Piding | WWTP Piding | 9,000 | TZW, from 03/2023 LGL (Erlangen) | 11/2020 - 12/2024 |
| Teisendorf | WWTP Teisendorf | 8,100 | TZW, from 03/2023 LGL (Erlangen) | 11/2020 - 12/2024 |

\*for both WWTPs in Nuremberg, Klärwerk I and Klärwerk II, operated in a network; with design capacities of 1,400,000 equivalent inhabitants for Klärwerk I and 230,000 equivalent inhabitants for Klärwerk II

At the inlet of the WWTPs, 24-hour composite samples were collected using autosamplers, either after grit removal or following mechanical treatment. In the Munich city district, sampling initially consisted of 3-hour composite samples collected between 6:00 and 9:00 in the morning, before being switched to 24-hour composite samples in March 2023. In Stadtbergen, qualified grab samples (QGS) were taken over the course of one hour, with six samples collected every 10 minutes in the morning. In Königsbrunn, 24-hour composite samples were collected using an autosampler, while QGS served as a backup method in case of sampler malfunction.

After sampling, the samples were transported to the corresponding laboratories at 4°C in less than 24 hours (Medema et al. 2020b; Markt et al., 2021). Samples were processed at the laboratory of the Technical University of Munich (TUM), the DVGW-Technologiezentrum Wasser (TZW), the laboratory at the Augsburg WWTP, and two laboratories of the Bavarian Health and Food Safety Authority (LGL) located in the city of Erlangen and municipality of Oberschleißheim (Table 1).

At the Augsburg WWTP, samples were collected at the WWTP, stored at +4°C, and extracted the following day using the Wizard Enviro TNA Kit (Promega) according to the manufacturer’s protocol. The SARS-CoV-2 genes N1 and N2 were subsequently quantified as biomarkers.

At the TUM, TZW and LGL laboratories, the viral RNA was extracted immediately after arrival and subsequently used to quantify several SARS-CoV-2 genes as biomarkers. The detailed extraction and quantification protocols from TUM and TZW are described in Mitranescu et al. (2022) and Ho et al. (2022).

At both laboratories of the LGL, a volume of approximately 45 ml wastewater of each sample was centrifuged at 4,500 g for 30 min to separate solid particles. For each sample, 40 ml supernatant was transferred to a new 50 ml tube and mixed with 10% PEG 8000 and 2.25% NaCl (w/v). After 1 h incubation on a rotator (60 rpm) at 4°C, the samples were centrifuged at 16,000 g for 1 h at 4°C. The resulting pellet was resuspended in 200 µl nuclease-free, PCR-grade water. For RNA extraction, automated sample processing was applied using Maxwell® RSC extraction instrument (Promega) (Oberschleißheim) or KingFisher™ Flex (Thermo Scientific™) (Erlangen) following the manufacturer’s recommendations. The samples were eluted in 50 – 100 µl.

In the different laboratories, up to four SARS-CoV-2 genes (selected from N1, N2, E, ORF1b, and RdRP2) were monitored in addition to the surrogate fecal indicators Pepper Mild Mottle Virus (PMMoV) and crAss-like phage (crAssphage) using either digital PCR at TUM (QIAcuity OneStep Advanced Probe Kit, Qiagen), digital-droplet PCR at TZW and both laboratories of the LGL (One-step RT-ddPCR Advanced Kit, BioRad), or RT-qPCR at Augsburg WWTP (GoTaq® Enviro Wastewater SARS-CoV-2 System, N1 and N2, Promega).

### Biomarker data processing

Laboratory and sampling data were recorded by the sampling and laboratory staff using ArcGIS Online (an ArcGIS Online license is required for access), the WebGIS cloud solution from Esri Deutschland GmbH, via the ArcGIS Survey123 native application for iOS, Android or Windows devices. This application served as a data entry interface, and all collected data were subsequently processed in an automated workflow (Figure 1). Clinical data, specifically individual SARS-CoV-2 case reports, was stored on local computers at the respective health authorities. To ensure data protection, these case reports were aggregated at the sewer service area level before being reported via ArcGIS Online. Only the aggregated, anonymized data was stored on the ArcGIS server, while the individual case data remained exclusively with the local health authorities to ensure privacy compliance, as illustrated in Figure 2.

**Figure 1:**
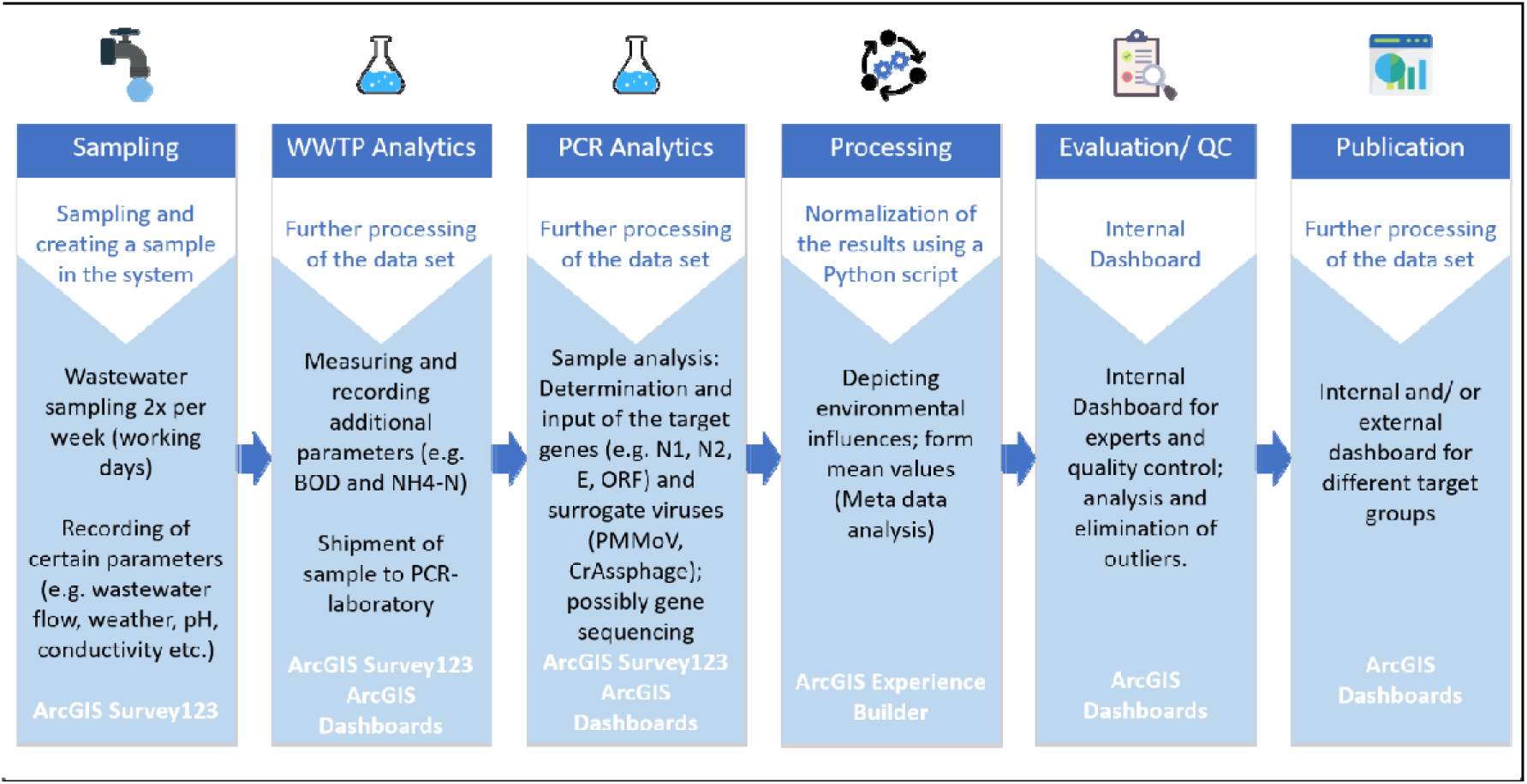
Workflow for sample processing and data presentation in the ANNA-WES data model

**Figure 2:**
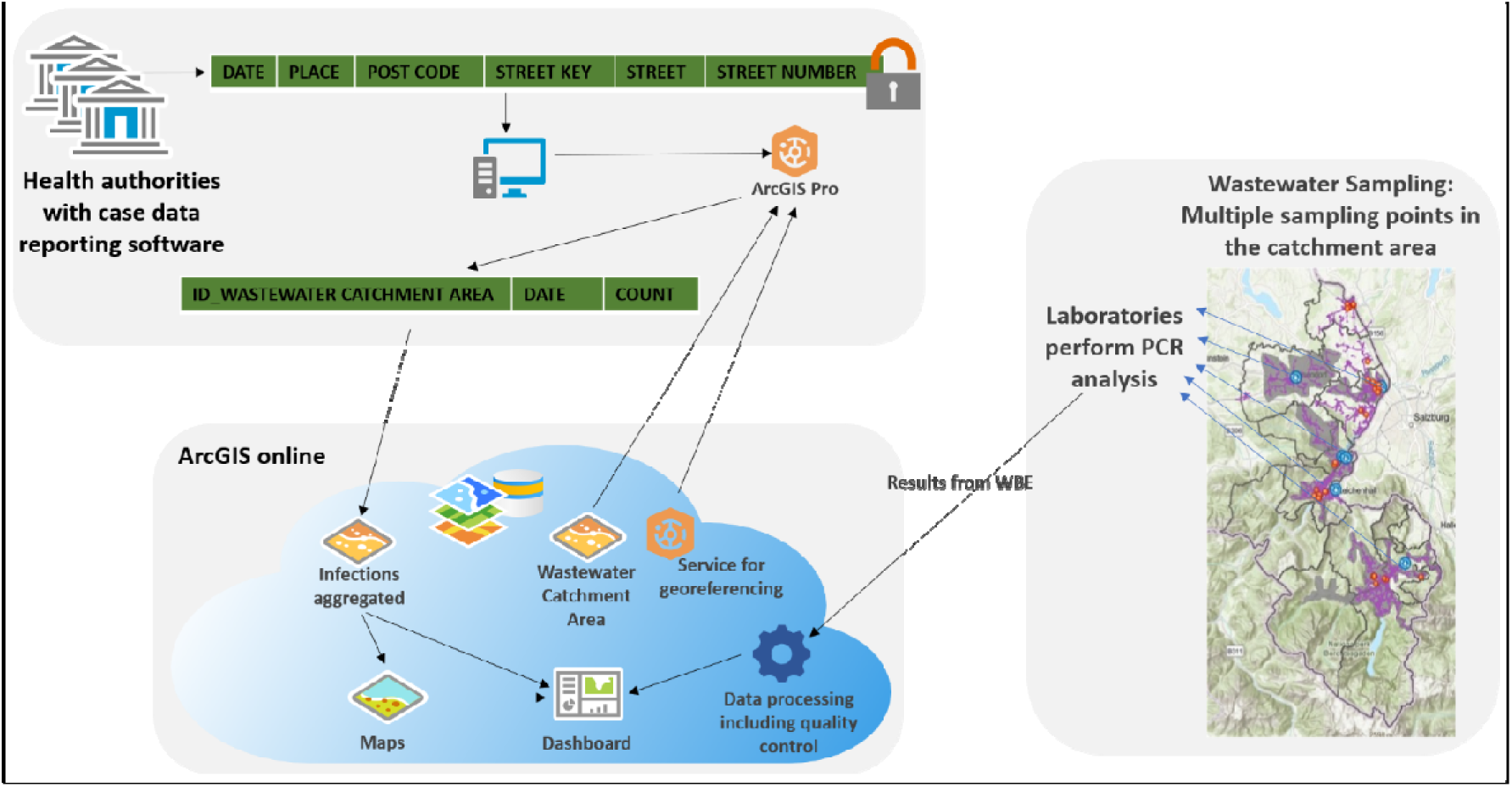
Transmission and storage of clinical case numbers and wastewater-based epidemiology results

The underlying data model was designed using UML in Microsoft Visio, created in ArcGIS Pro, and published to ArcGIS Online. It consists of multiple classes, with the measurement class containing all attributes of a sample, as listed below. Additional classes store information on other relevant information, e.g., sampling locations, laboratories, and sewer networks.

The sampling data included essential metadata such as the sampling date and time, sample type (composite or QGS), wastewater volume flow, and meteorological conditions (e.g., temperature, precipitation). Additional wastewater parameters, such as pH, wastewater temperature, conductivity, ammonia concentrations, and chemical oxygen demand (COD), were recorded when available. Laboratory data comprised biomarker concentrations obtained from PCR analyses, along with predefined LOQ and limits of detection (LOD) specific to each laboratory. The required data and sources for the ANNA-WES data model are listed in Supplement 2. To ensure traceability, a unique sample barcode was generated for each sample, and the PCR laboratory appended the biomarker copy numbers to the same dataset. Besides laboratory results and sampling metadata, a commentary section in ArcGIS Survey123 allowed the users to document any unusual events, such as sampler malfunctions or transport issues (e.g. samples arriving frozen). These samples were subsequently flagged as outliers and manually reviewed. If deemed to be true outliers, they were excluded from further data processing. This review process was implemented before the introduction of automated QC.

A data processing step was integrated using a Python script (see Supplement S3 for the script and Supplement S4 for the corresponding flowchart), which performed initial filtering, averaging, and normalization. The script was integrated within the ArcGIS Online data structure via ArcGIS Notebooks, allowing it to run automatically each night.

The following data handling procedure applies to cases where all target genes are above the laboratory-defined limit of quantification (LOQ) for the PCR analysis (Supplement S4). If one or more target genes fall below the detection limit, a placeholder value – half the LOQ as defined by the respective laboratory – is assigned to these undetectable genes to calculate the mean target gene value. The script then calculates the mean value of the biomarker genes and normalizes it based on either the wastewater volume flow (default for WWTPs with accurate flow data) or a mixed fecal indicator (MFI) derived from the surrogate viruses crAssphage and PMMoV (for samples lacking accurate flow data) (Mitranescu et al., 2022). If the required normalization parameter is missing for a given sampling location, the corresponding data point is excluded from further processing. No imputation is performed, and normalization is only applied when complete data is available. Samples with entries in the commentary section are automatically flagged for manual verification. If no comments are present, the running average over three sampling days is calculated from the normalized mean target gene value. This running average helps smooth short-term fluctuations in SARS-CoV-2 biomarkers and improves the visualization of weekly trends. The calculation follows a centered approach, meaning the value represents the mean of the previous, current, and following sampling dates. Additionally, alternative regression methods such as LOESS can be integrated (LGL, 2023; RKI, 2023a).

As the final step, the processed data is displayed in a user-friendly, customizable dashboard with clear trend indicators, designed to be easily understood by non-professionals and adaptable to the needs of each community. Additionally, dashboards were created to support data aggregation at higher levels, such as state or federal levels. The entire process, including data visualization, was governed by a rights and roles concept, ensuring that access to sampling sites and data was restricted based on user roles.

Access was granted through predefined view permissions, ensuring that users could only view or edit data relevant to their role. For example, wastewater operators have been assigned view-only access rights to data from their designated sites, whereas public health officials could both view and edit data across all relevant sites. This secure and flexible structure enables sample information to be accessed at any time, allowing authorized users to exclude data points as outliers from further processing, review, and potentially reinclude data points flagged in the commentary section, as well as make necessary corrections, such as fixing typos. However, in the initial phase of the project, these modifications were not automatically documented, which could introduce a potential source of bias – whether intended or unintended. To address this, we later developed and implemented an automated QC algorithm, which systematically flags outliers and applies standardized criteria for data validation. The algorithm and its implementation are described in the following section.

### Automated quality control and outlier detection

The automated QC algorithm, developed in Python (https://github.com/agblum/ssqn), provides an alternative data processing procedure to the general workflow presented above. A schematic diagram and a detailed explanation of the QC algorithm are presented in Supplement S5 and S6 and are published in the GitHub repository. In addition to the data processing steps mentioned above, the algorithm includes a data filtering step to remove outliers. It operates as a complete workflow, applied to an ArcGIS data table (or a similar CSV or XLS file with identical header names) as input for data filtering and normalization.

This algorithm enables automated quality filtering and categorization of data points across tables from multiple sites. It also includes a feature to flag cases requiring manual inspection, such as changes in gene ratios (see below). The script iterates through all data and metadata entered by wastewater operators and PCR laboratories, distinguishing between “outliers” (definitive outliers) and “flags” (putative outliers). By default, accumulating three putative outliers results in a definitive outlier. Entries in the comments field are automatically flagged as putative outliers.

The algorithm screens the following parameters using two to five statistical outlier detection methods: LOF (Local Outlier Factor), RF (Random Forest), IQR (Interquartile Range), Z-score, and CI (Confidence Interval). For an outlier to be confirmed, at least two methods must align, with LOF and IQR set as the default methods.

The parameters screened by the algorithm include:

(a) The ratio of individual SARS-CoV-2 genes, which are assumed to be constant. If three consecutive gene ratio outliers are detected, a manual inspection is required to check for potential viral mutations in the primer/probe sequences.
(b) The surrogate virus quantity.
(c) The daily flow, where a fixed (but adjustable) threshold of three times the dry weather flow is used instead of statistical outlier detection.
(d) Electrical conductivity and, if available, ammonia values.
(e) The seven-day reproduction value (R-value) (based on the Robert Koch-Institute (RKI) R-value (an der Heiden, Matthias (2023)); Similar to daily flow, fixed thresholds of 3.8 or 1/3.8 are used, based on literature values (an der Heiden, 2020; RKI, 2021), after prior normalization with the flow data.

All results and outlier sources are retained in plots and tables for manual inspection and further processing. The algorithm’s settings, as described above, are conservatively chosen. However, all thresholds and parameters can be adjusted as needed.

### GIS-based intersection of public health and wastewater-based data

Political boundaries, typically used for reporting public health data, often do not align with sewer service areas (see example Figure 6, city of Augsburg). Using GIS systems, we aligned clinical health data with sewer service areas, allowing clinical prevalence data – such as COVID-19 cases, testing and vaccination rates, and occupancy of intensive care units – to be aggregated and displayed based on wastewater service areas via the ArcGIS Experience Builder (Figure 2). To enable this intersection, we used polygon-based service definitions (ArcGIS Create Features: Polygon), derived from digitized versions of the sewer system typically available through local authorities or manually defined. The results from WBE are extracted from the database described above. Clinical datawas provided by local health authorities through their reporting software. These case reports were aggregated, anonymized, and centrally stored to comply with data protection regulations (Figure 2). Additionally, clinical COVID-19 infection data aligned with political jurisdiction boundaries was obtained from the Robert Koch Institute (RKI), Germany’s federal public health institute, responsible for monitoring and controlling infectious diseases. Because each patient’s post code, street key, street, and street number were included, we were able to intersect each patient’s registration address with the digital sewer service area, which ultimately allowed us to evaluate only the clinical data that fell within our polygonally defined sewer service area.

This alignment enables precise linkage of clinical data to sewer service areas, ensuring that both data sets reflect the same population within the same geographic area. Additionally, this approach allows us to extract clinical data specific to smaller areas, such as a city district, when sampling is conducted within localized areas (e.g., in a district sewer) rather than at a central WWTP.

To analyze whether aligning clinical case data to sewer service areas improves the correlation with wastewater biomarkers, we calculated Spearman’s rank correlation coefficients. Additionally, we performed a time-lag analysis, assuming that wastewater signals precede clinical case reports. To avoid potential biases introduced by different normalization methods, we used unnormalized bi-omarker concentrations for this analysis.

## 3. Results

In this study, we advanced the digitization of WES in close collaboration with public health authorities, developing ANNA-WES – an end-to-end model encompassing the entire process from sample collection to public reporting. Georeferenced dashboard websites with different designs were established for the counties of Berchtesgaden (Figure 3) and Augsburg (Supplement 7), enabling near realtime reporting of wastewater-based results. Both counties made these dashboards publicly accessible (Landratsamt Berchtesgadener Land, 2022; Presse Augsburg, 2023). Initially developed for monitoring SARS-CoV-2 at the local level, where seamless integration with public health structures with real-time data displays was achieved, this model was later expanded to the state level for the Free State of Bavaria (https://www.bay-voc.lmu.de/abwassermonitoring) and to the national level for Germany (RKI, 2023c).

**Figure 3:**
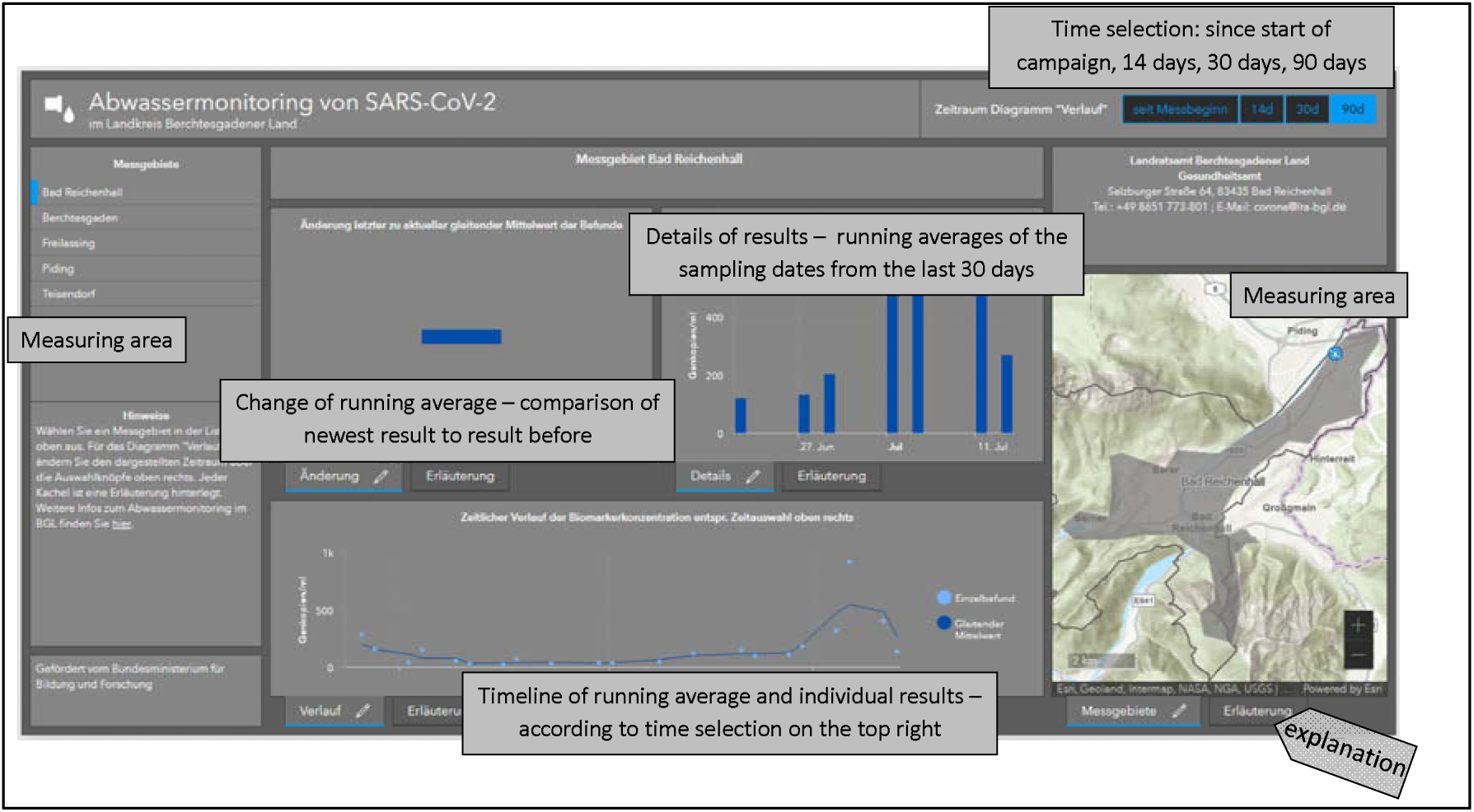
Design of the dashboard for SARS-CoV-2 wastewater-based epidemiology for the county of Berchtesgadener Land

Our implementation model includes:

– a GIS-based database,
– automated quality filters and controls,
– push notifications for out-of-specification events during sampling, transport, and laboratory procedures,
– georeferencing of public health and wastewater-based data
– data security and privacy measures, including anonymization and restricted access levels to sensitive health information,
– interoperability with other public health data systems for streamlined data sharing and analysis,
– expandability for biomarker and metadata (e.g., new sampling sites, sequencing data for variants, new biomarkers, further clinical data from public health departments),
– presentation of results through public and internal dashboards.

### Development and implementation of a real-time workflow and dashboard system for wastewater-based epidemiology

The workflow model shown in Figure 1 was initially developed and implemented during the COVID-19 pandemic to provide immediate support to collaborating health authorities. The workflow comprised the following steps:

(a) Creating new sample entries in the central GIS database and adding the corresponding wastewater parameters.
(b) Entering raw quantitative biomarker results for SARS-CoV-2 genes and surrogate biomarkers.
(c) Processing data via a Python script that averages, normalizes (Mitranescu et al., 2022), and smoothens the biomarker values.
(d) Allowing qualified users to view and edit input data to ensure data quality, as the automated QC algorithm had not yet been developed.
(e) Displaying the processed data on a digital dashboard in a clear and interpretable format.

As noted earlier, automated QC was not included in the initial stages of the project. Instead, manual review and editing served as preliminary QC measures, enabling the identification of key factors that later informed the development of an automated QC algorithm. This algorithm, now available for integration into the workflow, is described in the next chapter.

We designed a customized ArcGIS Survey123 application to minimize false reporting, which permitted laboratory entries only for preregistered samples submitted by wastewater operators. Clear instructions for data entry were provided for the input screens (Survey123 App for staff of WWTPs and webpage entry forms for staff of analytical laboratories). To support the initial establishment of the workflow, we conducted online workshops and introduced a “hotline” for assistance. These measures were crucial when rolling out the system to multiple sites, such as at the national level, which was implemented in 2022 when the data structure was made available to the German Ministry of Health (RKI, 2023c). The dashboard design for ANNA-WES was inspired by existing clinical ArcGIS COVID-19 dashboards, which facilitated the interpretation of results and made them comparable to the clinical dashboards used during the pandemic. A Frequently Asked Questions (FAQ) section on wastewater-based monitoring was provided for each element of the dashboard (see Supplement S17).

We designed two versions of the dashboard with different levels of detail. The first dashboard was a detailed dashboard intended for restricted use by the crisis management team, QC instances, and research. This dashboard included comprehensive data stored in an ArcGIS table format, such as gene copy numbers for each SARS-CoV-2 gene, aggregated clinical COVID-19 cases within WWTP service areas, and an editing option for qualified users to manually control data quality. Since automated QC was not yet available, users could edit or exclude data to address typos, sampling or laboratory errors, and outliers caused by unexpected events (e.g., heavy rain or frozen samples). Relevant metadata on wastewater parameters, weather conditions, and surrogate viruses was also collected to support data verification.

The second version was a simplified dashboard for the public domain, displaying only the essential information needed to interpret the wastewater data: the normalized, average concentration of SARS-CoV-2 biomarkers for a location over the whole sampling period displayed as single measurement points (dots) and the rolling average over three samples (line), the running biomarker concentration over the last 30 days, and the current trend (rising, constant, or declining SARS-CoV-2 biomarker concentration compared to the last 3 measurements), as shown in Figure 3. This dashboard was designed for integration into a website, as established at two of our sampling sites. In early 2023, the display of the results was consolidated and transferred from local authorities at the county level to the state-wide monitoring platform BayVOC (LGL, 2024), managed by LGL. The workflow was adapted to include new sampling sites and information, with the dashboard design revised to currently display monitoring data from 30 Bavarian cities, which is publicly accessible under https://www.bay-voc.lmu.de/abwassermonitoring (last accessed January 2025).

### Development of an automated quality control and outlier detection algorithm

As we gained experience and a deeper understanding of wastewater monitoring data, sewer system behavior, and the relationship between wastewater and clinical data, we developed a workflow to enable unsupervised data processing and outlier detection. Parameters relevant to wastewater-based monitoring, such as flow, ammonia, and conductivity, are integrated (with fully adjustable settings that allow its application to different conditions of wastewater systems and laboratory workflows), with normalization and plotting routines for visual outlier inspection. The algorithm was tested using data sets from nine different locations and four different laboratory workflows (see Table 1 and Supplement 1 for details on the sampling period and the number of data points tested) as it is known that there are systematic differences between laboratories (Wilhelm et al., 2023).

In the dataset from the city of Augsburg, measured at the Augsburg WWTP using RT-qPCR, no outliers were detected, likely due to the short sampling period of less than one year. In contrast, the QC steps identified specific outliers for inspection at all other locations and the city of Augsburg measured with dPCR (Figure 4A, Figures for the locations Munich, Nuremberg, Karlsruhe, Königsbrunn, Berchtesgaden, Ebersberg, and Piding see Supplements S8-S14). As shown in Figure 4B, these outliers appeared to be specific to the respective sites and laboratories. The highest frequency of outliers was due to the reproduction factors in communities with a low number of connected inhabitants (Königsbrunn, Berchtesgaden, Ebersberg, and Piding, all under 30,000 connected inhabitants), which apparently can lead to sudden peaks.

**Figure 4:**
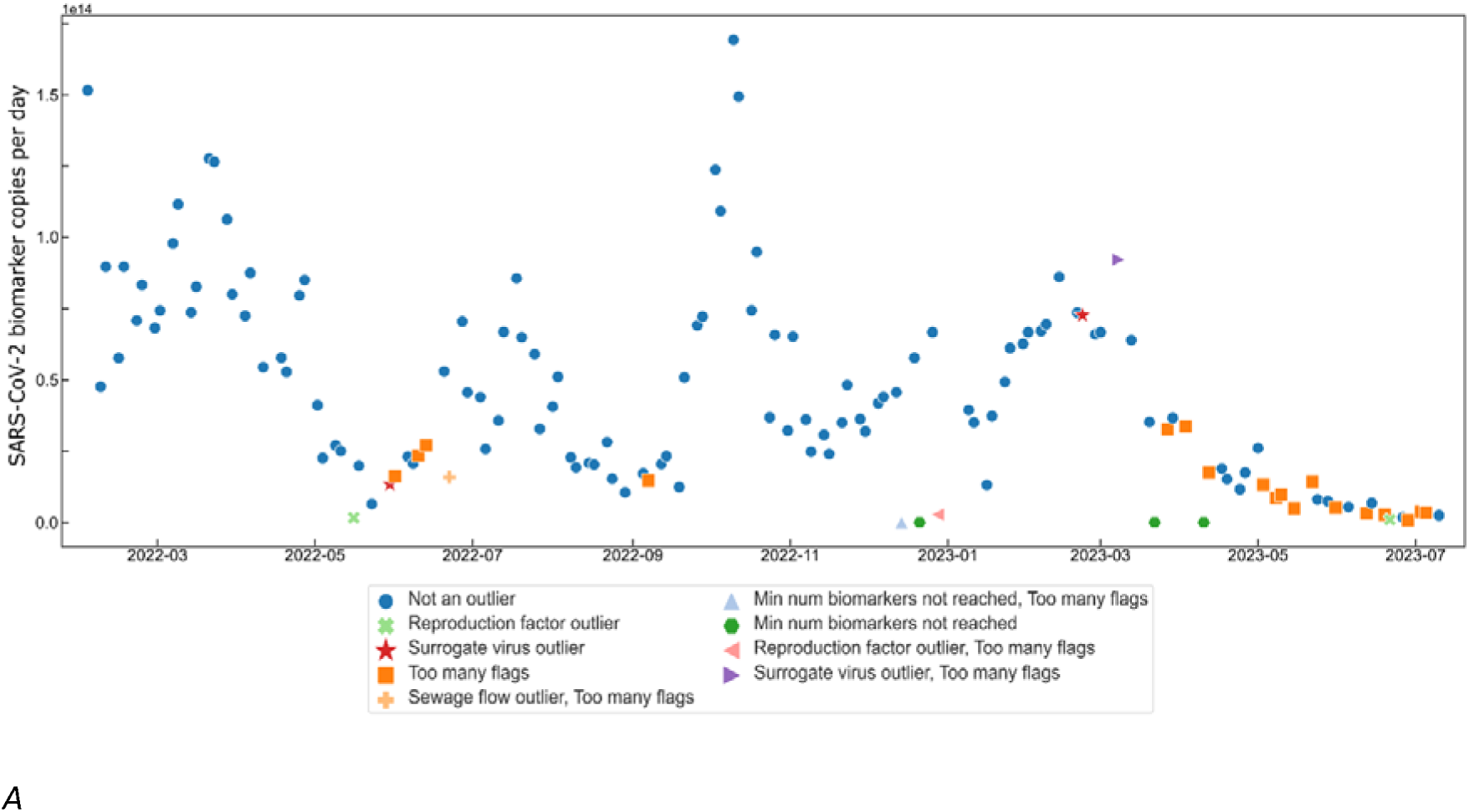
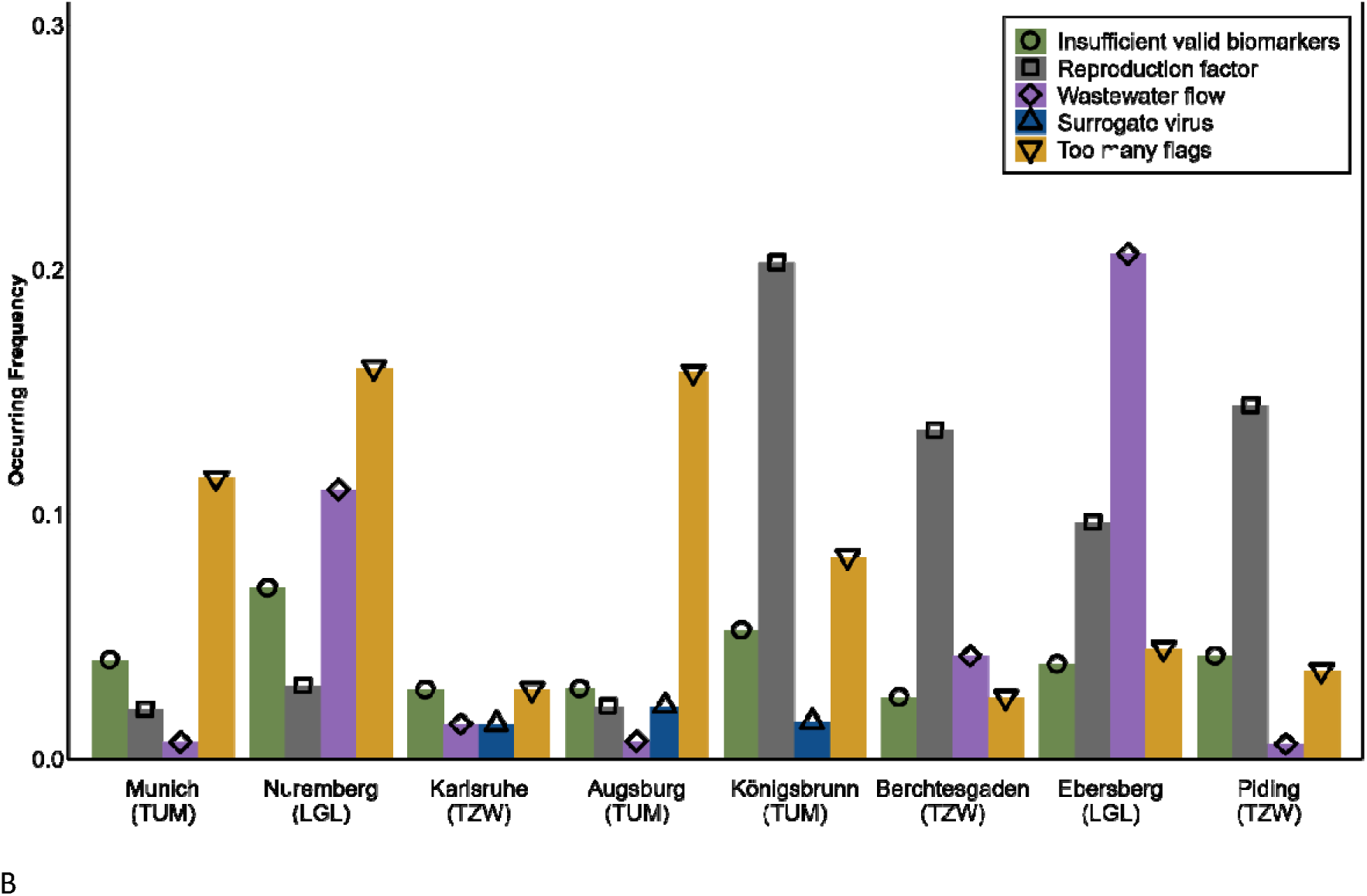
Outlier detection algorithm for a SARS-CoV-2 wastewater monitoring data set. A. Output of SARS-CoV-2 biomarker results for city of Augsburg (digital PCR) with outliers marked by the quality control algorithm with flow-normalized SARS-CoV-2 values. Detection of various outliers ranging from wastewater parameters (flow), number and ratio of detection genes (min number of biomarkers), surrogate viral parameters, too many irregularities (too many flags), or an outlier that is unrealistic from an epidemiological perspective (reproduction factor outlier). B. Frequency of outlier types by location, arranged by population size served (from highest to lowest), showing site-specific frequencies. Laboratory names are indicated in parentheses. The empirical technical error, estimated as the mean relative standard deviation (RSD)of the method is 14%.

While implemented for SARS-CoV-2 wastewater monitoring, the algorithm can be extended with an automated feedback loop that alerts the project manager when out-of-specification events, such as unusual gene ratios, are detected. These irregularities may also indicate the emergence of new viral variants with mutations in the primer binding sites, potentially necessitating adjustments in laboratory procedures.

### Alignment of public health data with wastewater service areas using GIS

The GIS system aligned public health data with the wastewater service area. This approach allowed us to assess whether the correlation between clinical and wastewater-based data improves when clinical data is matched to the population connected by a particular city-wide sewer system rather than to political boundaries, as clinical data was typically provided and reported based on these political boundaries during the pandemic. Clinical health data is typically strictly protected due to its sensitive and personal nature. Processing such data necessitated internal aggregation (Figure 2) to ensure anonymization and compliance with data protection regulations. We used ArcGIS Online to establish georeferenced cases to agglomerate and visualize the COVID-19 cases for the service areas of the communities sampled during this study (see examples for Munich and Augsburg in Figure 5 and 6). The GIS-corrected data can be used whenever comparisons of the wastewater data with public health data are required and results in accurate public health data that matches the wastewater-based data.

**Figure 5:**
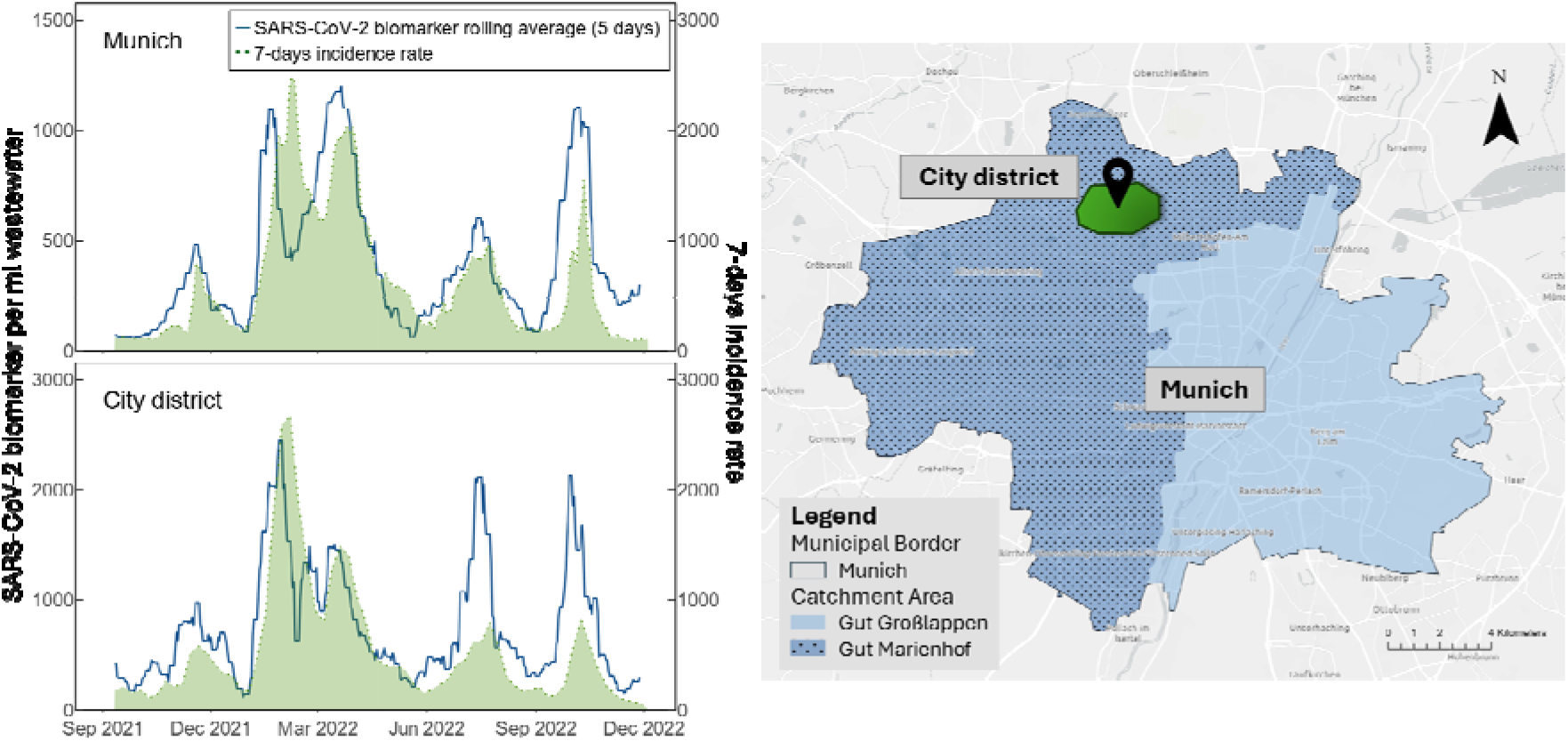
Map of the municipal border of the city of Munich with the wastewater service areas of its two wastewater treatment plants Gut Großlappen and Gut Marienhof, and a fictitious city district that is an example of the district we sampled. The graph on the left shows the 7-day incidence of the city of Munich and the SARS-CoV-2 biomarker counts of wastewater at the wastewater treatment plant Munich-Gut Großlappen and the city district.

**Figure 6:**
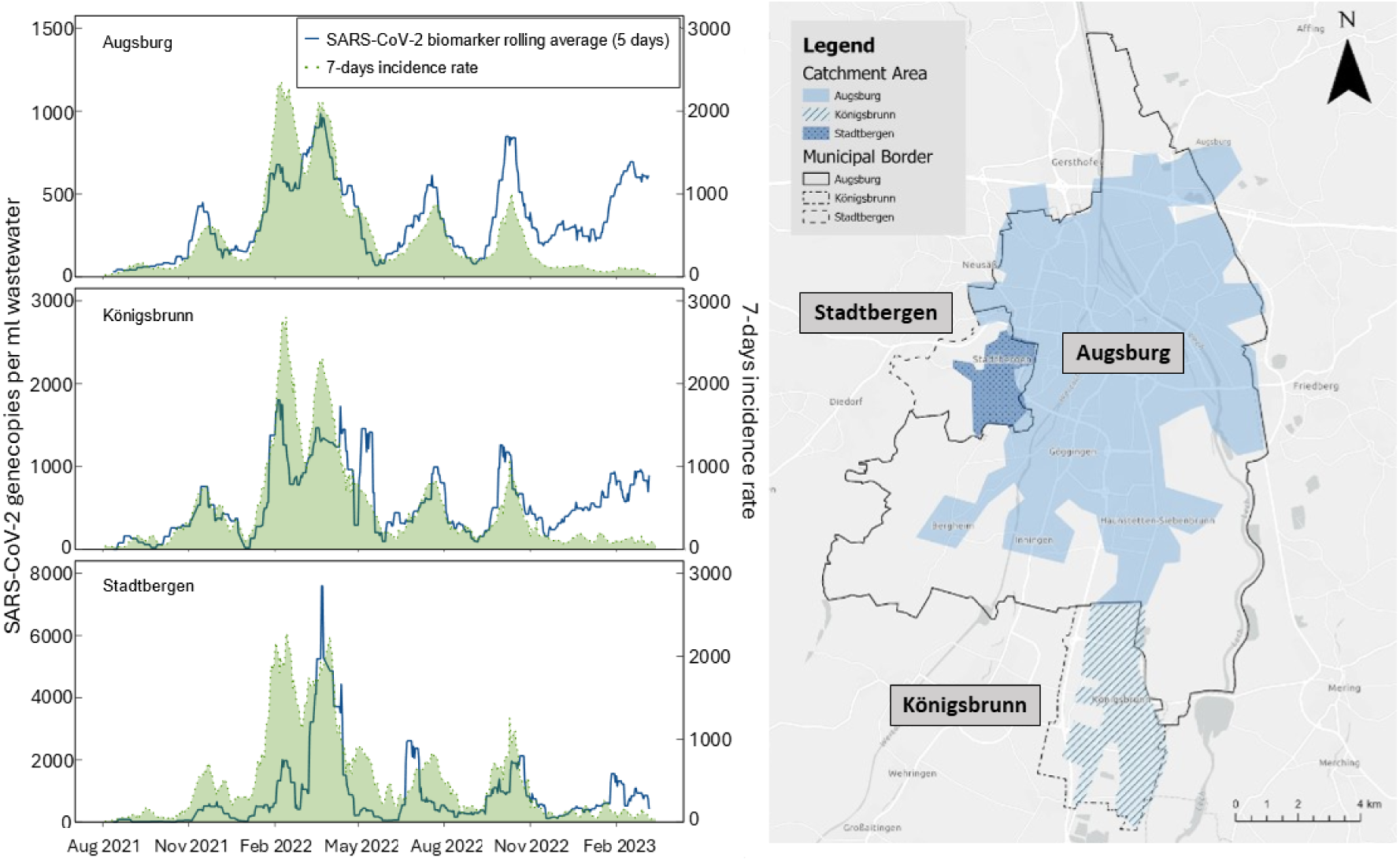
Difference between the municipal border and the service area of the WWTP of the city of Augsburg (Germany). Georeferenced case numbers can be calculated using a GIS system to obtain case numbers for the WWTP service area.

One of our sampling sites, the city of Munich, is served by two WWTPs: Gut Großlappen (2 million p.e.) and Gut Marienhof (1 million p.e.). In this study, wastewater sampling was conducted exclusively at the WWTP Munich-Gut Großlappen. Figure 5 illustrates the approximate service areas of the two wastewater plants under dry weather conditions, as well as a conceptual view of the sampled city district. Due to the dynamic operation of Munich’s sewer system, these service areas can shift during stormwater events. Smaller counties in the vicinity of Munich, which are also connected to the city’s sewer system, account for only a small proportion of the population and are not included here.

We sampled at WWTP Munich-Gut Großlappen, the larger of the two, which serves approximately 70 percent of the city’s population. As a result, the population represented in the infection data from the Robert-Koch Institute (RKI), which typically covers the entire city, differs from the population within the service area of the wastewater samples. Additionally, we collected samples from a city district (anonymized) serviced by a separate sewer system and a dedicated pump station. While samples from the WWTP were 24-hour composite samples, those from the city district were 3-hour composite samples taken during the morning.

We received anonymized SARS-CoV-2 infection data, extracted with GIS, corresponding to the service areas of WWTP Munich-Gut Großlappen and the city district from the city’s health department (Gesundheitsreferat Munich). We used Spearman’s rank-order correlation to quantitatively assess how well the extracted infection data matches the wastewater biomarker data compared to the infection data for the entire city of Munich provided by the RKI. Figure 5 shows the trajectory of the extracted infection data as 7-day incidences versus the corresponding SARS-CoV-2 biomarker concentrations from the wastewater measurements (not normalized, presented as a rolling average for 5 samples) covering the period from September 2021 to December 2022. The Spearman correlation analysis showed a strong overall correlation between clinical (both extracted and RKI) and wastewater biomarker data. Biomarker data from both the district and the full-scale WWTP revealed a slightly stronger correlation with the GIS-extracted infection data for the service areas than with the RKI infection data, except for the Omicron BA.5 and BQ variants occurring in the district. Table 2 provides the Spearman’s correlation coefficients (r) for the different SARS-CoV-2 variants.

**Table 2:** Spearman’s correlation coefficients between SARS-CoV-2 biomarkers from Munich wastewater treatment plant Munich-Gut Großlappen and the Munich city district, and the 7-day incidences for the extracted WWTP service area and Munich RKI incidences. The data covers different time periods from September 2021 to December 2022 and is grouped by dominant variants. Significance levels for the Spearman’s correlation coefficients are indicated as follows: *** for p < 0.001, ** for p < 0.01, and * for p < 0.05.

| SARS-CoV-2 biomarker | 7-days Incidence | Delta | Omicron |  |  |  | Whole period |
| --- | --- | --- | --- | --- | --- | --- | --- |
|  |  |  | BA.1 | BA.2 | BA.4 and BA.5 | BA.5 and BQ |  |
| WWTP | WWTP (GIS-extracted) | 0.90*** | 0.12 | 0.96*** | 0.94*** | 0.71*** | 0.81*** |
|  | Munich (RKI) | 0.82*** | 0.05 | 0.91*** | 0.88*** | 0.69*** | 0.78*** |
| District | District (GIS-extracted) | 0.71*** | 0.62*** | 0.93*** | 0.79*** | 0.80*** | 0.79*** |
|  | Munich (RKI) | 0.62*** | 0.43*** | 0.91*** | 0.78*** | 0.88*** | 0.77*** |

Since wastewater data often precedes clinical case data, (Medema et al., 2020a; Larsen & Wigginton, 2020; Ho et al., 2022), we performed an additional Spearman correlation analysis using varying time lags, with clinical data lagging behind the wastewater data by up to 15 days. Two main factors contribute to this temporal lead: individuals typically begin shedding the virus into wastewater before developing symptoms or seeking testing, and the wastewater testing and reporting pipeline is generally faster than the clinical reporting system. Additional variation arises from differences in shedding dynamics across variants and pathogens, sampling scheme, as well as from local sewer system characteristics.

Applying a time lag improved the correlations between clinical and wastewater data for all variants. The most pronounced effect was observed for Omicron BA.1, where a time lag of 13 days resulted in a correlation of r = 0.83*** (0.12 without timelag) for the WWTP service area and r = 0.73*** (0.05 without timelag) for RKI data with the Munich WWTP biomarker data, which is most likely attributed to its narrow peak abundance. A comprehensive table containing all Spearman’s correlation coefficients across variants and time lags is provided in Supplement S15.

We further evaluated the strength of the correlation between biomarker data and GIS-extracted infection data compared to the RKI data for another region in Southern Bavaria. Therefore, we sampled the influent of the WWTP of the city of Augsburg along with sewer sampling of two communities located in the county of Augsburg, Stadtbergen and Königsbrunn, both of which are sending their wastewater to the WWTP Augsburg. We collected 24-hour composite samples at the WWTP and Königsbrunn, while at Stadtbergen, we took QGS. When the autosampler was unavailable in Königsbrunn, we also received QGS samples. We obtained extracted health data from the local health department of the county of Augsburg. Figure 6 highlights the wastewater service area of the three sampling areas, illustrating the significant differences between the municipal boundaries and the actual sewer service areas. The corresponding biomarker concentrations and GIS-extracted clinical data are shown for each area.

The Spearman’s correlation analysis (Table 3) showed again a strong correlation between SARS-CoV-2 biomarkers and clinical incidence data for all dominant variants up to December 2022. However, from December 2022 onwards, the decline in clinical testing led to an underrepresentation of actual infection numbers, resulting in unreliable and weakened correlations beyond this period. A time lag analysis was also performed for these sampling sites, the results are displayed in Supplement S16.

**Table 3:**
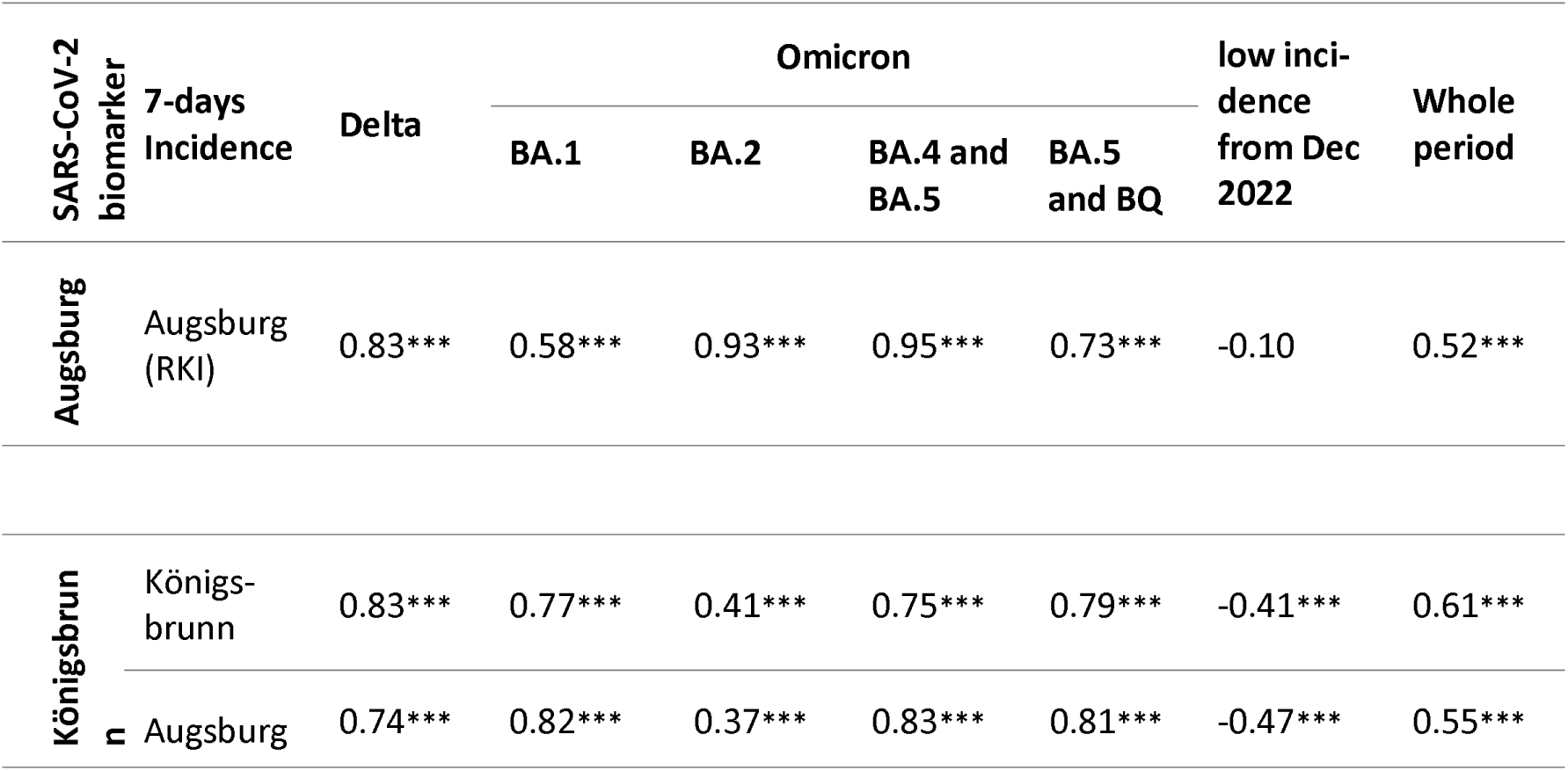
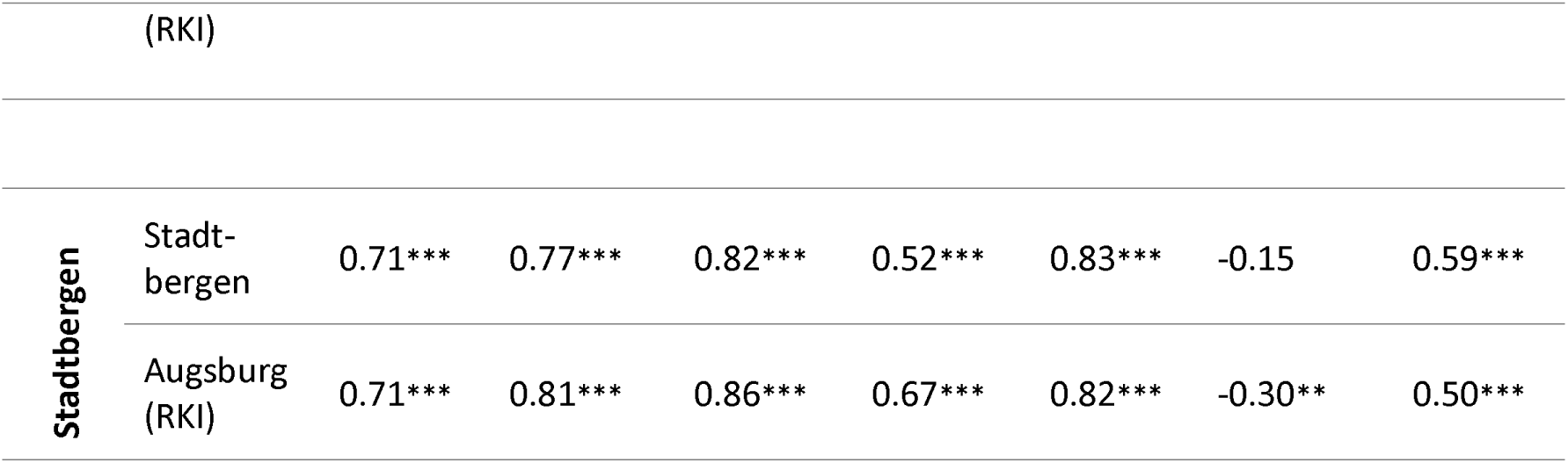
Spearman’s correlation coefficients of SARS-CoV-2 biomarkers of city of Augsburg and municipalities within the county of Augsburg (Königsbrunn and Stadtbergen) and 7-days incidences of the extracted WWTP service area and the Augsburg RKI incidences. Significance levels for the Spearman’s correlation coefficients are indicated as follows: *** for p < 0.001, ** for p < 0.01, and * for p < 0.05.

Despite the overall strong correlation, the Spearman analysis also revealed that incidence data from local health authorities, aligned to the sewer service areas, did not consistently correlate better than the RKI infection data aligned with political jurisdiction boundaries. This suggests that the sampling strategy, particularly the use of QGS, had a greater influence on the results than the spatial alignment of wastewater service areas. QGS capture only a brief snapshot of the day, unlike 24-hour composite samples that provide a more representative daily profile. Despite consistent sampling windows during morning hours, the natural variability in QGS samples seems to lead to greater fluctuations in biomarker concentrations compared to 24-hour samples. The increased variance in QGS samples limits the ability to assess the impact of geo-matching the wastewater service area. This emphasizes the need for harmonized sampling procedures and consistent use of composite samples to ensure data reliability. Nevertheless, the overall high correlations with clinical data suggest that wastewater analysis reliably reflects the infection trends and demonstrates the significant potential of WBE for monitoring smaller areas, such as villages and city districts.

Additionally, the GIS-based extraction and alignment of data proved valuable beyond correlation analysis. It facilitated the georeferenced visualization of results in a dashboard, allowing health authorities and the public to monitor incidence rates and wastewater biomarker concentrations specific to their areas of interest. During the pandemic, our collaborating health authorities benefited from this targeted approach, which enabled region-specific monitoring and enhanced their decision-making capabilities. This clearly demonstrates the added value of integrating GIS methods into WES workflows, supporting more effective public health responses.

## 4. Discussion

### Decentralized integration of wastewater-based data in a global context

Health authorities are often untrained in interpreting WBE data, yet their role in implementing disease containment measures makes understanding WBE results essential. From the onset of the COVID-19 pandemic, we collaborated closely with health authorities to develop user-friendly, accessible data presentations tailored for both professionals and the general public. Our workflow was codeveloped with local health authorities and wastewater treatment teams in the counties of Berchtesgadener Land and Augsburg, focusing on minimizing uncertainties in data entry and interpretation. Following validation and positive feedback, the data pipeline and dashboard were integrated into the national pilot project ESI-CorA, funded by the European Union (RKI, 2023c). Designed for scalability, the workflow expanded to include additional sampling sites with minimal adaptation. Recognizing the impracticality of providing in-depth training to all health authorities, we developed FAQ documents to facilitate understanding and accessibility (see FAQ in Supplement S17).

Furthermore, we supported the integration of the harmonized pipeline into the statewide WBE program BayVOC of the State of Bavaria (LGL, 2024). The pipeline has since been expanded to include additional communities and is now featured in an enhanced, publicly accessible dashboard, with results reported within a maximum of 48 hours. Building on these foundations, we developed an automated QC algorithm as a separate feature to further improve data reliability and reproducibility. While the QC algorithm is still undergoing long-term and real-time testing, the harmonized workflow and data processing have demonstrated scalability and adaptability. It is noteworthy that the transfer of responsibility for managing wastewater monitoring entries and data handling from the leading university (TUM) to the Bavarian Ministry of Health proved successful, highlighting the potential for transferring these processes from research initiatives to public health authorities.

### Current limitations and the importance of robust and accessible data for public health decision-making

To guide public health decision-making, accurate and robust epidemiological, clinical, and laboratory data must be available and accessible early in an epidemic. Publicly accessible dashboards and visualizations are often positively received by both the public, health authorities, and political decision makers (Dixon et al., 2022). Consistent, georeferenced epidemiological data is crucial for understanding the transmissibility, geographic spread, transmission routes, and risk factors of infections. This information provides a baseline for epidemiological modeling, aiding authorities in planning response efforts and containment strategies to reduce the disease burden. Moreover, real-time access to detailed data is critical for prioritizing surveillance efforts (Dong et al., 2020). Effective local crisis management, supported by real-time local dashboards – whether publicly available or confidential – is essential for an accurate crisis situation assessment and subsequent decision-making (Focosi et al., 2024; Stoney et al., 2023). These dashboards enable local authorities to respond more quickly, precisely, and effectively to the specific circumstances in their area. Additionally, they contribute to a more effective national or transboundary pandemic management for broader policy decisions.

However, significant challenges remain regarding the accessibility and sharing of public health data. Although there are global commitments to share such data, a lack of systematic frameworks, harmonized data-sharing models, and cohesive federal and global guidelines persists (Gigerenzer et al., 2022; van Panhuis et al., 2014). Naughton et al. (2023a) identified numerous discrepancies in the presentation and accessibility of the wastewater monitoring solutions, often addressed on a case-by-case basis. Given that WBE involves multiple stakeholders – such as WWTPs, laboratories, health authorities, and government bodies – establishing a harmonized data flow with automated transmission and QC is essential for better pandemic preparedness within the public health sector (Roßmann et al., 2022; McNabb, 2010; EU, 2021). Without harmonized approaches, variability in data normalization, analysis, and visualization on dashboards could impair the understanding of infection data across different sites, ultimately affecting crisis management (Naughton et al., 2023a).

### Quality control in wastewater-based epidemiology

As valuable as a dashboard might be for assessing and comparing a pandemic situation, the quality of the data fed into the system remains crucial. Our experience has shown that significant quality differences can arise from varying sampling strategies (Bertels et al., 2022, Uchaikina et al. 2023), sample preparation protocols (Wilhelm et al., 2023), and qPCR and dPCR methods employed (Nyaruaba et al., 2022; Wilhelm et al., 2023). To ensure accurate and comparable data, it is crucial to test and establish standardized operating procedures (SOPs) for both sampling and laboratory processes (Kitajima et al., 2020; Keshaviah et al., 2021; Ciannella et al., 2023; Keenum et al., 2024). However, other factors across WBE workflows also influence data quality.

Our QC algorithm bridges a critical gap by systematically assessing data reliability through statistical outlier detection and integration of epidemiological and wastewater parameters. While previous efforts focused on minimizing inter-laboratory variability (van Nuijs et al., 2018; Ahmed & Bivins et al., 2020; Kantor et al., 2021; Wilhelm et al., 2023), reliable data must accommodate diverse laboratory workflows. The QC algorithm extends beyond laboratory-based approaches by systematically identifying anomalies – such as typos, extraordinary values, and unusual gene ratios – and flagging entries for manual inspection. While the QC currently aims to address the main type of outliers, it cannot address all sources of variation. Instead, it enables the detection of unexpected patterns or deviations that may result from the catchment, varying protocols, instruments, or sampling strategies. The QC is designed to be robust across different laboratory conditions, but we recommend that interlaboratory cross-validation studies be conducted where quantitative comparability between sites is a primary goal. This would complement the QC approach by providing additional assurance of analytical consistency, particularly in multicenter surveillance programs. Future versions of the QC may incorporate the expected variation for specific laboratory workflows.

The algorithm’s modular design allows for adjustable thresholds, enabling its application across diverse wastewater systems, laboratory workflows, and application to new biomarkers. Thresholds for outlier detection (e.g., Z-score, LOF, IQR) were selected based on established practices in environmental and epidemiological data analysis and refined through empirical testing with our dataset and evaluation. While no formal sensitivity or Receiver Operating Characteristic (ROC) analysis was conducted, the chosen thresholds were intentionally conservative to minimize false positives and support robust anomaly detection across heterogeneous data sources. We recommend that future studies systematically evaluate threshold performance using sensitivity analyses, such as ROC curves, particularly when applying the algorithm to new biomarkers or datasets. This flexibility of the algorithm’s modular structure and threshold settings was evidenced by its successful deployment across multiple locations with varying sample collection methods and laboratory protocols. Additionally, the algorithm alerts project managers to potential viral mutations in primer or probe binding sites, enabling timely laboratory adjustments. This scalable solution enhances data reliability across laboratories, particularly in large-scale WBE programs.

Beyond laboratory QC, wastewater data variability is heavily influenced by sewer system factors. Normalization techniques, such as using flow rates, human biomarkers, or wastewater parameters as correction factors, are commonly employed to mitigate these effects. However, these methods often fail to account for all influencing factors, and their effectiveness varies across communities, with some cases even degrading data quality (Mitranescu et al., 2022; O’Brien et al., 2023). Smoothing algorithms, such as rolling averages or LOESS regression, are another common approach to stabilize trends (Arabzadeh et al., 2023). While effective for reducing noise, these methods do not address the root causes of variability.

To overcome these limitations, our QC algorithm incorporates controls for flow rates, electrical conductivity, and ammonia levels, effectively identifying and excluding anomalies caused by high wet-weather events, industrial discharges, or sewer system dynamics. For example, high-flow events dilute wastewater and dislodge biofilms, introducing biases. Additionally, we integrate the R-reproduction value as an independent parameter, ensuring wastewater results align with theoretical epidemic trends and do not exceed expected daily changes. By combining these controls with adaptable normalization techniques and embedded smoothing algorithms, the QC algorithm enhances trend detection and data accuracy, providing a more robust framework for managing variability in WBE.

Eventually, further investigation into the complexities of sewer systems remains necessary. Although our QC algorithm accounts for various factors, influences from sewer systems on biomarker results remain a “black box” in our understanding (Bertels et al., 2022). The outliers identified by the algorithms are partially explainable by, e.g., wet-weather events or variations in the laboratory procedures. However, for example, positive outliers that were detected by the R-value filtering must be due to other reasons that require further investigation. We speculate that factors such as wet-weather events, flow rate variations, industrial discharge, physico-chemical properties of wastewater, biofilm retention, flow surges, and travel time within the sewer network can introduce noise and lead to unexpected, potentially delayed spikes or dips in biomarker concentrations. Moreover, population dynamics, such as commuting patterns and large public events, likely influence biomarker concentrations (Schmid et. al, 2024). In addition to potential intrinsic normalization methods, cell phone data may be used in the future (Rhodes et al., 2024) to further calibrate the connected population. It has been shown that this can further enhance the precision of the wastewater monitoring (Schenk et al., 2024).

The automated quality control algorithm relies on static thresholds for outlier detection. While effective, these thresholds may not adapt to changing conditions or emerging trends in wastewater data, for instance new variants emerged. Integrating machine learning (ML) approaches could possibly enhance adaptability by dynamically adjusting thresholds based on historical data and metadata. ML has already demonstrated success in epidemiology and environmental monitoring, offering opportunities to improve both outlier detection and the interpretation of underlying variability in wastewater data (Wiens & Shenoy, 2018; Wu et al., 2018; Barapatre et al., 2023; Liu et al., 2023; Zehnder et al., 2023). However, challenges such as model interpretability, data protection compliance, and the need for continuous retraining must be carefully addressed. By leveraging ML, future QC algorithms could improve data reliability and support real-time, context-aware decision-making, bridging the gap between data collection and actionable public health insights.

### GIS integration and flexible data workflows: benefits and future implications for wastewater-based epidemiology

In the future, WBE is expected to expand as a surveillance and early warning system for other diseases and biomarkers (EU, 2021; European Commission, 2022). It is now embedded in the revised EU Urban Wastewater Treatment Directive (EU UWWTD) (EU, 2024) and in some communities, WBE is already being used to monitor antibiotic resistance (Larsson et al., 2023; Stange et al., 2023) and respiratory diseases like influenza (APA-OTS, 2023; LGL, 2024).

The ArcGIS database offers flexibility by allowing the addition of new biomarkers or SARS-CoV-2 variants through simple modifications to the structured input tables. This enables the creation of additional dashboards with consistent design logic, supporting the inclusion of new pathogens, laboratories, sampling sites, and user-specific access levels for institutions and regions. The geo-based data storage makes our workflow particularly suitable for communities where sewer service areas differ significantly from political boundaries or for distinct areas within a city, as it allows health data to be aligned with actual catchment areas. While aligning public health data with sewer service areas may not be critical for routine monitoring programs approximating the community by the sewer system (Tscharke et al., 2019), it is essential when precise intersections are needed, e.g., for statistical analysis.

An additional advantage of a GIS-based platform is its ability to anonymize public health data while preserving critical information. This is particularly important when sampling smaller areas, such as a street via a sewer access point, to protect individual privacy and maintain public trust in health surveillance methods (Emam et al., 2015). The GIS structure of our data pipeline facilitates the secure collection and anonymization of data, enabling detailed sharing with health authorities while presenting only anonymized results to the public. Notably, sensitive information, such as clinical data, is stored locally within health institutions, with only anonymized data transferred to the GIS cloud, ensuring strict compliance with health data protection regulations.

Nevertheless, reliance on commercial GIS software (ArcGIS Online, Survey123, and GIS dashboards) presents constraints for broader adoption, particularly in regions without existing licenses or infrastructure. While many municipalities already work with ArcGIS licenses, this dependency can hinder adoption in regions lacking the necessary infrastructure or resources to obtain and use such software. Future efforts should focus on developing an open-source implementation for GIS data aligned with WHO and EU recommendations that propose to use the PHES-ODM, an open data structure for WES (https://github.com/Big-Life-Lab/PHES-ODM). However, while PHES-ODM offers a valuable foundation for harmonized data architecture, it currently lacks integrated features for automated quality control, near real-time reporting, or practical alignment with public health service areas via GIS. By integrating georeferenced data, automated QC algorithms, and Python-based normalization and visualization scripts, ANNA-WES closes this gap. It provides a ready-to-use, field-tested workflow designed not only for interoperability but also for direct application in health authority decision-making. This makes ANNA-WES a complementary and implementation-ready extension to existing open data frameworks.

Uniformity is particularly crucial when introducing new biomarkers that require calibration. During the pandemic, we calibrated WBE using COVID-19 clinical case data, but future biomarkers will likely necessitate further calibration with public health data (Sloan et al., 2022).

Given that the comprehensive evaluation of georeferenced public health data is often restricted to public authorities (Peters & Zeeb, 2022), a potential future scenario could involve establishing reference sites – cities where GIS-based public health systems are developed. These systems would allow future WES markers to be calibrated against various public health parameters or socio-economic factors, improving the accuracy and applicability of WES data. Additionally, integrating other environmental data (e.g., remote sensing) into a GIS-based database would enable the construction of comprehensive dashboards that consider both public and environmental health within the broader One Health framework (Singer et al., 2023).

### Perspectives

We successfully applied ANNA-WES across multiple sites in Germany, demonstrating its adaptability to different laboratory workflows and sampling strategies. However, broader validation across international regions is essential to confirm its robustness under diverse infrastructural and environmental conditions. Factors such as rain events and industrial discharges can strongly affect biomarker levels and vary by region. While our QC algorithm accounts for some of these influences, their underlying mechanisms remain insufficiently understood. Future research is needed to better mitigate their impact and enhance the interpretation of WBE trends.

Beyond technical feasibility, sustainable integration of WBE into routine public health practice remains a key challenge. Transferring responsibility from academic initiatives to health authorities requires coordinated implementation, staffing, and training. Many public health agencies lack experience with environmental or GIS-based data, underscoring the need for structured onboarding and ongoing support. Our success relied on regular stakeholder communication and accessible guidance materials (e.g., FAQs, explainer charts). Interagency working groups can support long-term knowledge exchange and harmonize implementation. While this manuscript focuses on technical development, future efforts should aim to establish practical guidelines and foster intersectoral coordination.

ANNA-WES is designed for scalability and flexibility, making it a valuable tool even in endemic scenarios. Its modular framework is not limited to SARS-CoV-2 and can incorporate additional biomarkers and environmental indicators. By leveraging existing local infrastructure, the system enables costeffective deployment, even in resource-constrained settings.

## 5. Conclusion

In the current transition from pandemic to endemic infections, where public willingness to undergo testing has decreased, and health data has become fragmented, WES has emerged as a reliable source for capturing information across the entire population. Harmonized strategies for data sharing, quality control, and presentation are crucial to prevent and manage pandemics. Our adaptable framework marks a vital step forward in improving public health surveillance.

Our advancements, including an automated workflow, a GIS-based database, tailored solutions for diverse sewer service areas, and a QC algorithm, streamline WES programs for all stakeholders, such as health authorities, wastewater teams, and data analysts. The immediate display of wastewater monitoring results, with the option to expand to other pathogens and scale up to regional, national, or global dashboards, makes this system highly flexible and adaptable. It supports interdisciplinary collaboration and accelerates communication, which is especially valuable in times of overloaded healthcare systems during pandemics. Integrating WBE and public health data into widely used GIS systems, the ANNA-WES model is a versatile decision-support tool for managing pandemics, urban planning, and environmental surveillance. By incorporating this system into routine monitoring programs, we can ensure a faster, more coordinated response to future pandemics and potentially prevent future outbreaks through early health monitoring and preventive measures.

The successful implementation of WES requires not only national and international collaboration but also close interdisciplinary cooperation between health and environmental authorities. By working together, these sectors can ensure more accurate, holistic data analysis and response strategies that address both public health and environmental factors influencing disease transmission. Cross-sector collaboration will enable a more integrated approach to managing future pandemics and environmental health challenges.

While the workflow and QC algorithm are critical first steps toward harmonizing WES data processing, ensuring readiness for future pandemics requires immediate integration of these systems and the development of expertise. Securing funding for personnel across health authorities, laboratories, and WWTPs is equally important, as is establishing standardized operating procedures in databases. Further research is needed to better understand how sewer system factors impact WES results. These elements, along with strong interdisciplinary collaboration, and national and international cooperation, are vital to enhancing both our preparedness and response to future pandemics.

## Abbreviations

ANNA-WES: Automated Network for Normalization, Analysis, and Visualization of Wastewater and Environmental Surveillance
CI: Confidence Interval
COD: Chemical oxygen demand
COVID-19: Coronavirus disease 2019
GIS: Geographic information system
IQR: Interquartile Range
LOD: Limit of Detection
LOF: Local Outlier Factor
LOQ: Limit of Quantification
ML: Machine learning
PCR: Polymerase chain reaction
dPCR: Digital polymerase chain reaction
ddPCR: Digital-droplet polymerase chain reaction
qPCR: Quantitative polymerase chain reaction
RF: Random Forest
RT-qPCR: Quantitative reverse transcription polymerase chain reaction
SARS-CoV-2: Severe acute respiratory syndrome coronavirus 2
SSQN: SARS-CoV-2 sewage quality control and normalization
RKI: Robert Koch-Institute
RNA: Ribonucleic acid
ROC: Receiver Operating Characteristic
R-value: Reproduction value
QC: Quality control
QGS: Qualified grab samples
WBE: Wastewater-based epidemiology
WES: Wastewater and environmental surveillance
WHO: World Health Organization
WWTP: Wastewater treatment plant

## Acknowledgments

This study was financially supported by the German Federal Ministry of Education and Research as part of the funding program Sustainable Water Management (NaWaM-RiSKWa) (Biomarker, grant number 02WRS1557) and the Bavarian State Ministry of Health and Care (StMGP). Additional funding was provided through the AMELAG project, funded by the German Federal Ministry of Health (BMG). A. G. received funding the Bay-VOC project, funded by the StMGP. I.M. was a subcontractor by PTKA. C.Wb. was financed by DFG WU 890/2-1.

We sincerely thank the staff of the wastewater treatment plants in Augsburg, Bad Reichenhall, Berchtesgaden, Ebersberg, Freilassing, Karlsruhe, Munich, Nuremberg, Piding, Teisendorf, and Weiden for their invaluable support. We also extend our gratitude to the health authorities from the counties of Augsburg and Berchtesgadener Land, as well as Munich, for their close collaboration throughout the pandemic. We also wish to thank the Medical Intelligence & Information division of the Sanitätsakademie der Bundeswehr, especially Dr. Alexander Ziegler, for their continuous support and valuable insights, particularly during the challenging period of the pandemic and the early stages of the WBE program.

A special thank you goes to Carolin Kerscher, Lucia Maciossek, Heidrun Mayrhofer, Meenakshi Prasad, Dominik Kugler, Darshita Singal, Andreas Mirlach, Veronika Hillebrand, Naim Vilabrera, and Claus Lindenblatt, as well as other lab colleagues whose dedication, meticulous lab work, and tireless efforts were instrumental to this study. Their commitment ensured the reliability of sample processing, analysis, and transportation, and we deeply appreciate their hard work. Finally, we thank Elisa Schorer for her valuable support with GIS and database management. The graphical abstract was generated using OpenAI’s DALL·E tool. Final design and content were reviewed and approved by the authors.

## Data availability

The data for the individual dashboards is stored in the GIS system. The Python scripts for the metadata analysis and display of the results can be found in the supplementary material. The Python script for the quality control algorithm including example data from our sites can be found on GitHub under: https://github.com/agblum/ssqn. After the end of our project, the monitored municipalities were now included in the federal and governmental monitoring program of Bavaria (https://www.bay-voc.lmu.de/) and Germany (https://www.rki.de/DE/Content/Institut/OrgEinheiten/Abt3/FG32/Abwassersurveillance/Abwassersurveillance.html). The detailed data model objects and data structure that was developed by Esri Deutschland GmbH is available (in German) at TUM. A licensed version implemented in ArcGIS Online is currently handled by Projektträger Karlsruhe (PTKA).

## 6. Author contributions

**Anna Uchaikina**: Conceptualization, Data Curation, Methodology, Software, Validation, Formal Analysis, Investigation, Visualization, Writing - Original Draft, Writing - Review & Editing, Resources, Project Administration.

**Anna-Sonia Kau**: Conceptualization, Data Curation, Methodology, Software, Validation, Formal Analysis, Investigation, Visualization, Writing - Original Draft, Writing - Review & Editing, Resources, Project Administration.

**Alexander Graf**: Conceptualization, Data Curation, Methodology, Software, Validation, Formal Analysis, Investigation, Visualization, Writing - Review & Editing.

**Christine Walzik**: Conceptualization, Data Curation, Methodology, Software, Validation, Formal Analysis, Investigation, Visualization, Writing - Review & Editing, Resources.

**Alexander Mitranescu**: Conceptualization, Data Curation, Methodology, Software, Validation, Formal Analysis, Investigation, Visualization, Writing - Review & Editing.

**Lisa Falk**: Conceptualization, Data Curation, Methodology, Software, Validation, Formal Analysis, Investigation, Visualization, Writing - Review & Editing, Resources.

**Mohammad Shehryaar Khan**: Data Curation, Software, Validation, Formal Analysis, Investigation, Visualization, Writing - Review & Editing, Resources.

**Claudia Stange**: Conceptualization, Data Curation, Methodology, Validation, Formal Analysis, Investigation, Writing - Review & Editing, Resources.

**Johannes Ho**: Conceptualization, Data Curation, Methodology, Validation, Formal Analysis, Investigation, Writing - Review & Editing, Resources.

**Katalyn Roßmann**: Conceptualization, Data Curation, Methodology, Investigation, Writing - Review & Editing, Resources, Supervision.

**Ingo Michels**: Conceptualization, Data Curation, Methodology, Software, Investigation, Visualization, Writing - Review & Editing.

**Nathan Obermaier**: Data Curation, Methodology, Validation, Formal Analysis, Investigation, Writing - Review & Editing.

**Cristina J. Saravia**: Data Curation, Methodology, Validation, Formal Analysis, Investigation, Writing - Review & Editing.

**Susanne Rost**: Data Curation, Investigation, Visualization, Writing - Review & Editing, Resources.

**Thorsten Portain**: Data Curation, Investigation, Visualization, Writing - Review & Editing, Resources.

**Jürgen Demeter:** Data Curation, Methodology, Investigation, Writing - Review & Editing, Resources.

**Christopher Becker**: Data Curation, Methodology, Investigation, Writing - Review & Editing, Resources.

**Martina Füchsle**: Data Curation, Methodology, Investigation, Writing - Review & Editing, Resources.

**Fabienne Kaymaz-Ried**: Data Curation, Methodology, Investigation, Writing - Review & Editing, Resources.

**Alexander Klaus**: Data Curation, Investigation, Writing - Review & Editing, Resources.

**Tobias Ziegler**: Data Curation, Methodology, Investigation, Writing - Review & Editing, Resources.

**Katharina Springer**: Data Curation, Methodology, Investigation, Writing - Review & Editing, Resources.

**Melissa Hohl**: Data Curation, Methodology, Investigation, Writing - Review & Editing, Resources.

**Peter-Louis Plaumann**: Data Curation, Methodology, Investigation, Writing - Review & Editing, Resources.

**Annemarie Bschorer**: Data Curation, Methodology, Investigation, Writing - Review & Editing, Resources.

**Stefanie Huber**: Data Curation, Investigation, Writing - Review & Editing, Resources, Supervision.

**Patrick Dudler**: Data Curation, Investigation, Writing - Review & Editing, Resources, Supervision, Funding Acquisition.

**Andreas Tiehm**: Conceptualization, Data Curation, Methodology, Validation, Investigation, Writing - Review & Editing, Supervision, Funding Acquisition.

**Jörg E. Drewes**: Conceptualization, Data Curation, Methodology, Validation, Investigation, Visualization, Writing - Original Draft, Writing - Review & Editing, Resources, Project Administration, Supervision, Funding Acquisition.

**Christian Wurzbacher**: Conceptualization, Data Curation, Methodology, Software, Validation, Formal Analysis, Investigation, Visualization, Writing - Original Draft, Writing - Review & Editing, Resources, Project Administration, Supervision, Funding Acquisition.

