## Supplement for "From wastewater to GIS-based reporting: the ANNA-WES data model for reliable biomarker tracking in wastewater and environmental surveillance"

**From wastewater to GIS-derived reporting: a novel data model for biomarker tracking in wastewater-based epidemiology**

3: Bavarian Health and Food Safety Authority, Veterinärstraße 2, 85764 Oberschleißheim, Germany

4: Department of Water Microbiology, TZW: DVGW-Technologiezentrum Wasser, Karlsruher Straße 84, 76139 Karlsruhe, Germany

5: Medical Intelligence & Information (MI2), Surveillance & Multinationaler Force Health Protection Nexus, Sanitätsakademie der Bundeswehr, Munich, Germany

6: Esri Deutschland GmbH, Ringstraße 7, 85402 Kranzberg

7: Wastewater Technology Research, Wastewater Disposal, German Environment Agency, Corrensplatz 1, 14195 Berlin

8: Public Health Department, County of Augsburg, Prinzregentenplatz 4; 86150 Augsburg, Germany

9: Public Health Department, County of Berchtesgadener Land, Salzburger Straße 64; 83435 Bad Reichenhall, Germany

10: Public Works Department Augsburg, Klärwerkstraße 10, 86154 Augsburg, Germany

11: Health Departement of Munich, Gesundheitsreferat (GSR), Geschäftsbereich Gesundheitsplanung, Abteilung Daten und Evaluation, GSR-GP-DE, Bayerstr. 28a, 80335 München

### **Supplemental Information**

**List of Supplemental Text, Tables, and Figures**

**Supplement S1:** Detailed information on sampling locations (Table)

**Supplement S2:** Required Data Inputs and Sources for the ANNA-WES Data Model (Text)

**Supplement S3**: Data processing workflow for filtering, averaging, normalization, and 3 days rolling average calculation according to Mitranescu et al. (2022) (Text)

**Supplement S4:** Flow chart of the data processing workflow for filtering, averaging, normalization, and three days rolling average calculation according to Mitranescu et al. (2022) (Figure, created with Miro)

**Supplement S5:** Description of the quality control algorithm SSQN (Text)

**Supplement S6:** Flow chart of the quality control algorithm (Figure, created with Miro)

**Supplement S7:** Alternative design of the ANNA-WES dashboard for SARS-CoV-2 wastewater-based epidemiology created for the county of Augsburg (Figure)

**Supplement S8:** Outlier detection – Munich (Figure)

**Supplement S9:** Outlier detection – Nuremberg (Figure)

**Supplement S10:** Outlier detection – Karlsruhe (Figure)

**Supplement S11:** Outlier detection – Königsbrunn (Figure)

**Supplement S12:** Outlier detection – Berchtesgaden (Figure)

**Supplement S13:** Outlier detection – Ebersberg (Figure)

**Supplement S14:** Outlier detection – Piding (Figure)

**Supplement S15:** Spearman’s rank-order correlation analysis between wastewater-based epidemiology results and clinical severe acute respiratory syndrome coronavirus 2 (SARS-CoV-2) data for the city of Munich and a Munich city district, tested for various time lags across different SARS-CoV-2 variants and the entire sampling period. The time lag (in days) represents the lead time of wastewater data compared to clinical data, based on the assumption that wastewater data provide earlier results due to rapid sampling, laboratory processing, analysis, and reporting (significance: *** p < 0.001, ** p < 0.01, * p < 0.05). (Table)

**Supplement S16:** Spearman’s rank-order correlation analysis between wastewater-based epidemiology results and clinical severe acute respiratory syndrome coronavirus 2 (SARS-CoV-2) data for the city of Augsburg and the communities of Königsbrunn and Stadtbergen in the county of Augsburg, tested for various time lags across different SARS-CoV-2 variants and the entire sampling period. The time lag (in days) represents the lead time of wastewater data compared to clinical data, based on the assumption that wastewater data provide earlier results due to rapid sampling, laboratory processing, analysis, and reporting (significance: *** p < 0.001, ** p < 0.01, * p < 0.05).

**Supplement S17: Frequently Asked Questions (FAQ): How to use and interpret the ANNA-WES dashboard**

**Supplement S1:** Detailed information on sampling locations (Table)

| **Sampled community** | **Population served** | **Sampling site** | **Sample type** | **Sampling time** | **Laboratory** | **Sampling period** | **Sampling period  for QC** | **Sampling  days QC** |
| --- | --- | --- | --- | --- | --- | --- | --- | --- |
| Munich | 1,156,000 | WWTP Gut Großlappen | CS | 24 h | TUM | 07/2021 - 12/2024 | 02/2022 - 07/2023 | 148 |
| Nuremberg | 589,000* | WWTP Nuremberg Klärwerk I | CS | 24 h | LGL | 12/2022 - 12/2024 | 10/2023 - 11/2024 | 98 |
| Karlsruhe | 370,000 | WWTP Karlsruhe | CS | 24 h | TZW | 07/2020 - 12/2024 | 02/2022 - 06/2023 | 139 |
| Augsburg | 343,000 | WWTP Augsburg | CS | 24 h | TUM | 07/2021 - 12/2024 | 02/2022 - 07/2023 | 136 |
| Augsburg | 343,000 | WWTP Augsburg | CS | 24 h | Augsburg WWTP | 02/2022 - 07/2022 | 02/2022 - 07/2022 | 46 |
| Weiden | 43,400 | WWTP Weiden | CS | 17 h, later 24 h | TUM | 07/2021 - 12/2024 | - | - |
| Königsbrunn (Augsburg county) | 29,500 | Sewer manhole | CS** | 24 h | TUM | 07/2021 - 12/2024 | 02/2022 - 07/2023 | 131 |
| Berchtesgaden | 27,000 | WWTP Berchtesgaden | CS | 3.5 hours, later 24 hours | TZW, from 03/23 LGL | 07/2021 - 12/2024 | - | - |
| Munich district | 25,000 | Pump station | CS | 3 h, later 24 h | TUM | 07/2021 - 05/2023 | - | - |
| Freilassing | 20,000 | WWTP Freilassing | CS | 4 h, later 24 h | TZW, from 03/23 LGL | 07/2021 - 12/2024 | - | - |
| Bad Reichenhall | 19,500 | WWTP Piding | CS | 4 h, later 24 h | TZW, from 03/23 LGL | 07/2021 - 12/2024 | 02/2022 - 07/2023 | 116 |
| Stadtbergen (Augsburg county) | 15,000 | Sewer manhole | QGS | 6 samples every 10 min in the morning | TUM | 07/2021 - 12/2024 | - | - |
| Ebersberg | 12,500 | WWTP Ebersberg | CS | 24 h | LGL | 04/2023 - 12/2024 | 04/2023 - 11/2024 | 154 |
| Piding | 9,000 | WWTP Piding | CS | 4 h, later 24 h | TZW, from 03/23 LGL | 07/2021 - 12/2024 | 02/2022 - 11/2023 | 160 |
| Teisendorf | 8,100 | WWTP Teisendorf | CS | 4 h, later 24 h | TZW, from 03/23 LGL | 07/2021 - 12/2024 | - | - |
| CS = composite sample QGS = qualified grab sample QC = quality control | | *for both WWTPs in Nuremberg, Klärwerk I and Klärwerk II, operated in a network; with design capacities of 1,400,000 equivalent inhabitants for Klärwerk I and 230,000 equivalent inhabitants for Klärwerk II  **or QGS in case of sampler failure | | | | | | |

**Supplement S2:** Required Data Inputs and Sources for the ANNA-WES Data Model (Text)

Our developed data model ANNA-WES integrates several data sources to enable comprehensive analysis in wastewater-based epidemiology (WBE). This supplemental item lists all required and optional data parameters that must be entered via the application to facilitate accurate processing through the algorithm and effective visualization on the dashboard. Parameters marked in **bold** represent the minimal data inputs necessary for the process.

1) Wastewater parameters: **sample ID**, **operator ID**, **sampling date**, sewer overflow event, type of sample, **sampling period**, **sewage flow**, **dry weather condition**, temperature, conductivity, ammonia, type of sewage system, percentage of groundwater inflow. The sewage flow is recorded as the medium flow volume during the sampling period. Optionally recorded data at the WWTPs are pH-value, COD, NH_4_-N, conductivity, and shipping date of the sample to the lab. A field for additional **comments** is provided for notes about irregularities during sampling.

2) Laboratory parameters: **sample ID, laboratory ID, date of extraction**, **date of quantitative analysis,** **biomarker X gene copies**, **surrogate virus gene copies**, genome sequencing, VOC percentage, **limit of detection (X)**, and sample integrity. For data entry at the PCR lab, a **comment field** is also available for noting any irregularities. Once all obligatory data points are collected, the data undergoes processing.

3) Public health parameters: **location**, COVID-19 infections, **daily case data**, age statistics, testing rates, vaccination status, and hospitalization rates, **agglomerated georeferences**. During this project, this data was provided by the health authorities. During this project, these data were provided by the health authorities. Clinical prevalence data were supplied daily on workdays by the health department, with data from the weekend included in Monday's reports. The data were extracted in table format from the public health authority’s information software. A Python script created with ArcGIS Notebooks was employed to extract the data from the cloud daily, process it, and update the dashboards automatically.

4) Georeferenced data: Sampling site coordinates, **coordinates of the WWTP**, **georeferenced polygons** defining the wastewater catchment area. A digitized wastewater network can be used to infer the polygons.

The metadata from points 1 and 2 was organized into a large table, stored in an ArcGIS database, with parameters as columns and sample IDs as rows. This data structure was implemented in various database formats and could be exported as CSV or Excel files. The georeferenced data from points 3 and 4 was initially processed and securely stored on a local computer at the health authorities. To comply with data protection policies, the data was then aggregated and transferred to ArcGIS Online with restricted access.

**Supplement S3:** Data processing workflow for filtering, averaging, normalization, and 3 days rolling average calculation according to Mitranescu et al. (2022) (Text)

A detailed description of the Python script can be found in the SI of Mitranescu et al. (2022).

import numpy as np

import pandas as pd

\# %% import table

table_Augsburg = pd.read_excel('samples_Augsburg.xlsx')

\# %% 1) Reference time period for Internal Process Control: computing mean and std of log values for filter interval

table_Augsburg_reference_period = table_Augsburg.loc[(table_Augsburg['date_of_sampling'] <= '2022-05-11')]

\# 1.1) compute log10 values of surrogate viruses

table_Augsburg_reference_period['PMMoV_log'] = np.log10(table_Augsburg_reference_period['PMMoV_(copies/ml)'])

\# 1.2) calculate parameters (mean, standard deviation) for filter interval

mean_PMMoV_log_reference_period = np.mean(table_Augsburg_reference_period['PMMoV_log'])

std_PMMoV_log_reference_period = np.std(table_Augsburg_reference_period['PMMoV_log'])

\# 1.3) define filter intervals of PMMoV

lower_threshold_PMMoV_reference_period = mean_PMMoV_log_reference_period - 2 * std_PMMoV_log_reference_period

upper_threshold_PMMoV_reference_period = mean_PMMoV_log_reference_period + 2 * std_PMMoV_log_reference_period

\# %% 2) Internal Process Control by surrogate viruses (PMMoV)

\# 2.1) compute log10 values of surrogate viruses

table_Augsburg['PMMoV_log'] = np.log10(table_Augsburg['PMMoV_(copies/ml)'])

\# 2.2) PMMoV filter interval

table_Augsburg['PMMoV_Confidence_Value'] = np.nan

for i in range(table_Augsburg.shape[0]):

if (table_Augsburg['PMMoV_log'].iloc[i] < lower_threshold_PMMoV_reference_period) | (table_Augsburg['PMMoV_log'].iloc[i] > upper_threshold_PMMoV_reference_period):

table_Augsburg['PMMoV_Confidence_Value'].iloc[i] = False

else:

table_Augsburg['PMMoV_Confidence_Value'].iloc[i] = True

\# 2.3) remove days outside of filter interval --> set to PMMoV and genes to nan

for i in range(table_Augsburg.shape[0]):

if (table_Augsburg['PMMoV_Confidence_Value'].iloc[i] == False):

table_Augsburg['PMMoV_(copies/ml)'].iloc[i] = np.nan

table_Augsburg['N2_(copies/ml)'].iloc[i] = np.nan

table_Augsburg['ORF1_(copies/ml)'].iloc[i] = np.nan

table_Augsburg['E_(copies/ml)'].iloc[i] = np.nan

\# %% 3) Computation of average_genes

detection_limit = 1

\# 3.1) preallocate rows with nan in real table

table_Augsburg['average_genes'] = np.nan

table_Augsburg['average_genes_value_below_DL'] = np.nan

\# 3.2) define table and empty rows

table_Augsburg_average_genes_comp = table_Augsburg.loc[(pd.isnull(table_Augsburg['E_(copies/ml)']) == False)]

table_Augsburg_average_genes_comp['number_genes_below_DL'] = np.nan

table_Augsburg_average_genes_comp['average_genes_positive_finding'] = np.nan

table_Augsburg_average_genes_comp['average_genes'] = np.nan

table_Augsburg_average_genes_comp['average_genes_value_below_DL'] = np.nan

\# 3.3) fill rows with content

for i in range(table_Augsburg_average_genes_comp.shape[0]):

genes_below_DL = [(table_Augsburg_average_genes_comp['E_(copies/ml)'].iloc[i] < detection_limit),(table_Augsburg_average_genes_comp['ORF1_(copies/ml)'].iloc[i] < detection_limit),(table_Augsburg_average_genes_comp['N2_(copies/ml)'].iloc[i] < detection_limit)]

table_Augsburg_average_genes_comp['number_genes_below_DL'].iloc[i] = sum(genes_below_DL)

if table_Augsburg_average_genes_comp['number_genes_below_DL'].iloc[i] == 0:

table_Augsburg_average_genes_comp['average_genes_positive_finding'].iloc[i] = True

average_genes = (table_Augsburg_average_genes_comp['E_(copies/ml)'].iloc[i] + table_Augsburg_average_genes_comp['ORF1_(copies/ml)'].iloc[i] + table_Augsburg_average_genes_comp['N2_(copies/ml)'].iloc[i])/3

table_Augsburg_average_genes_comp['average_genes'].iloc[i] = average_genes

elif table_Augsburg_average_genes_comp['number_genes_below_DL'].iloc[i] == 1:

table_Augsburg_average_genes_comp['average_genes_positive_finding'].iloc[i] = True

sum_genes = 0

if table_Augsburg_average_genes_comp['E_(copies/ml)'].iloc[i] < detection_limit:

sum_genes = sum_genes + 0.5*detection_limit

elif table_Augsburg_average_genes_comp['E_(copies/ml)'].iloc[i] >= detection_limit:

sum_genes = sum_genes + table_Augsburg_average_genes_comp['E_(copies/ml)'].iloc[i]

if table_Augsburg_average_genes_comp['ORF1_(copies/ml)'].iloc[i] < detection_limit:

sum_genes = sum_genes + 0.5*detection_limit

elif table_Augsburg_average_genes_comp['ORF1_(copies/ml)'].iloc[i] >= detection_limit:

sum_genes = sum_genes + table_Augsburg_average_genes_comp['ORF_(copies/ml)'].iloc[i]

if table_Augsburg_average_genes_comp['N2_(copies/ml)'].iloc[i] < detection_limit:

sum_genes = sum_genes + 0.5*detection_limit

elif table_Augsburg_average_genes_comp['N2_(copies/ml)'].iloc[i] >= detection_limit:

sum_genes = sum_genes + table_Augsburg_average_genes_comp['N2_(copies/ml)'].iloc[i]

table_Augsburg_average_genes_comp['average_genes'].iloc[i] = sum_genes/3

elif table_Augsburg_average_genes_comp['number_genes_below_DL'].iloc[i] > 1:

table_Augsburg_average_genes_comp['average_genes_positive_finding'].iloc[i] = False

table_Augsburg_average_genes_comp['average_genes_value_below_DL'].iloc[i] = detection_limit/2

\# 3.4) retransfer average_genes in normal, long table

for i in range(table_Augsburg.shape[0]):

for j in range(table_Augsburg_average_genes_comp.shape[0]):

if table_Augsburg['date_of_sampling'].iloc[i] == table_Augsburg_average_genes_comp['date_of_sampling'].iloc[j]:

table_Augsburg['average_genes'].iloc[i] = table_Augsburg_average_genes_comp['average_genes'].iloc[j]

table_Augsburg['average_genes_value_below_DL'].iloc[i] = table_Augsburg_average_genes_comp['average_genes_value_below_DL'].iloc[j]

\# %% 4) Metadata Analysis

\# for days with operational issues, etc.:

\# omit "average_genes" values from "average_genes" column and save them in a separate "average_genes_metadata_flagged"-column

table_Augsburg['average_genes_metadata_flagged'] = np.nan

for i in range(table_Augsburg.shape[0]):

if (pd.isnull(table_Augsburg['comment_operation'].iloc[i]) == False):

table_Augsburg['average_genes_metadata_flagged'].iloc[i] = table_Augsburg['average_genes'].iloc[i]

table_Augsburg['average_genes'].iloc[i] = np.nan

\# %% 5) Normalization by flow

table_Augsburg['average_genes_norm_flow'] = table_Augsburg['average_genes']*table_Augsburg['flow_(qm/d)']/1e5

\# 1e5 is dry minimum determined by WWTP Augsburg

\# %% 6) Rolling Mean (3 days))

\# define short table, only with sample days

table_Augsburg_short = table_Augsburg.dropna(subset=['average_genes_norm_flow'])

\# create rolling mean

rolling_mean = table_Augsburg_short['average_genes_norm_flow'].rolling(3, min_periods=1, center = True).mean()

table_Augsburg_short['average_genes_norm_flow_mean'] = rolling_mean

rolling_mean = table_Augsburg_short['average_genes'].rolling(3, min_periods=1, center = True).mean()

table_Augsburg_short['average_genes_mean'] = rolling_mean

**Supplement S4:** Flow chart of the data processing workflow for filtering, averaging, normalization, and three days rolling average calculation according to Mitranescu et al. (2022) (Figure, created with Miro)

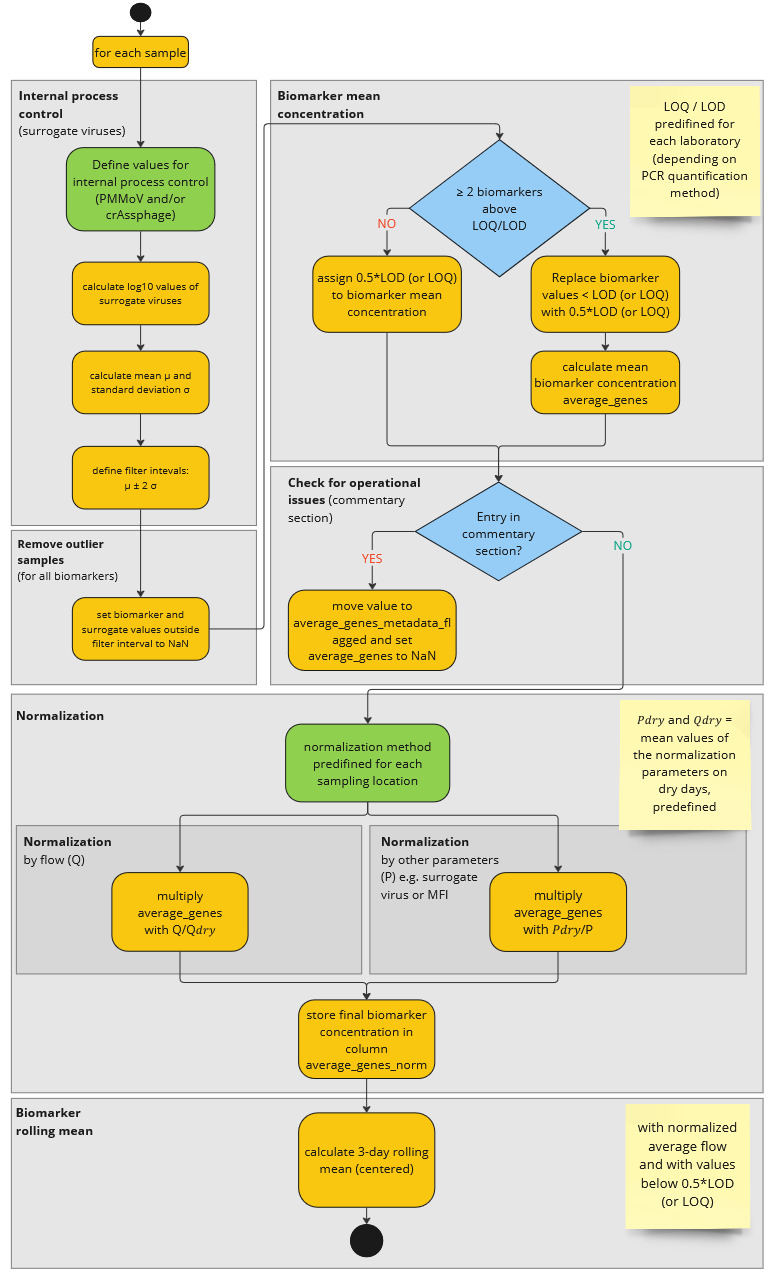

**Supplement S5:** Description of the quality control algorithm SSQN (Text)

The flow chart describes the quality control algorithm SSQN - SARS-CoV-2 sewage Quality control and Normalization available in Github: <https://github.com/agblum/ssqn>. The high-resolution flow chart can be found in the GitHub repository.

The algorithm processes the data in a stepwise manner (as indicated by the grey boxes), starting from the left:

a) **Plausibility**: Plausibility checks are applied to data table entries, evaluating the date, missing values, flow data, detection limits of biomarkers, the number of biomarkers, and the minimum number of measurements (n = 9).

b) **Biomarker Ratio**: The algorithm calculates pairwise ratios of target biomarkers (e.g., SARS-CoV-2 genes) where applicable and detects outliers in these ratios.

c) **Surrogates**: Surrogate markers (e.g., PMMoV and crAssphage) are screened for outliers.

d) **Sewage Flow**: Outliers in sewage flow are identified based on average dry weather conditions calculated from the dataset. Three different thresholds (factor 3, 2, and 1.5) are used to adjust the severity of the outlier detection.

e) **Water Quality**: Water quality parameters, such as conductivity and ammonia, are evaluated for outliers.

f) **Normalization and Reproduction Rate**: Biomarkers are normalized (in this case to sewage flow), and outliers in the reproduction rate are detected by considering previous measurements prior to the data point.

g) **Identification of Outliers**: At the end of the process, putative outliers identified in steps a) through f) are flagged. A flag counter is then applied to exclude data points with an excessive number of flags (default = 3).

**Supplement S6:** Flow chart of the quality control algorithm SSQN (Figure, created with Miro)

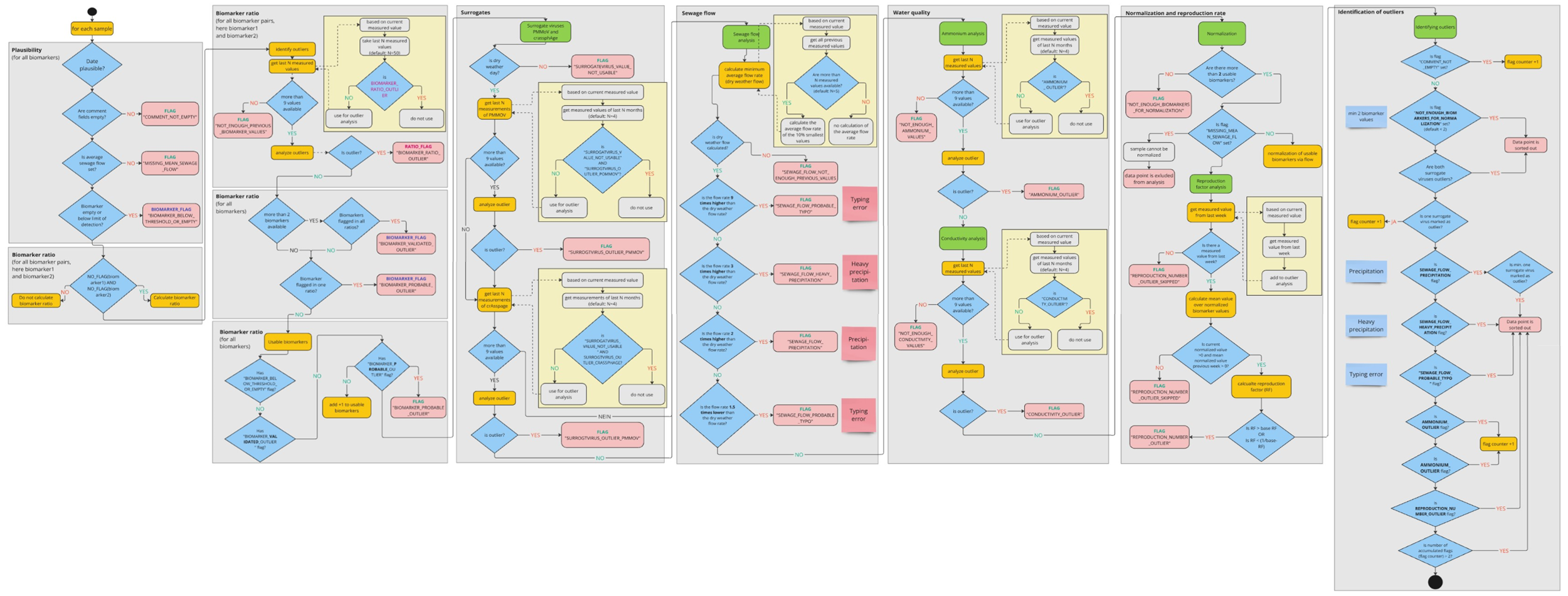

**Supplement S7:** Alternative design of the ANNA-WES dashboard for SARS-CoV-2 wastewater-based epidemiology created for the county of Augsburg (Figure)

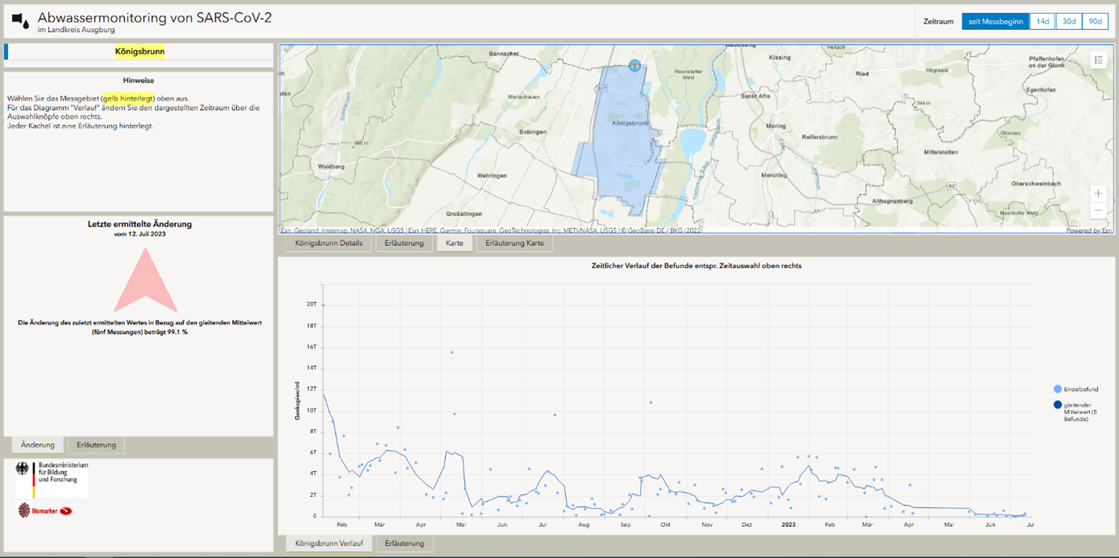

**Supplement S8 – S14:** Output of the outlier detection algorithm SSQN for SARS-CoV-2 wastewater monitoring data sets for Munich, Nuremberg, Karlsruhe, Königsbrunn, Berchtesgaden, Ebersberg, Piding. The outliers are marked by the quality control algorithm SSQN with sewage flow-normalized SARS-CoV-2 values. Detection of various outliers ranging from wastewater parameters (flow), number and ratio of detection genes (min number of biomarkers), surrogate viral parameters, too many irregularities (too many flags), or an outlier that is unrealistic from an epidemiological perspective (reproduction factor outlier).

**Supplement S8:** Outlier detection – Munich (Figure)

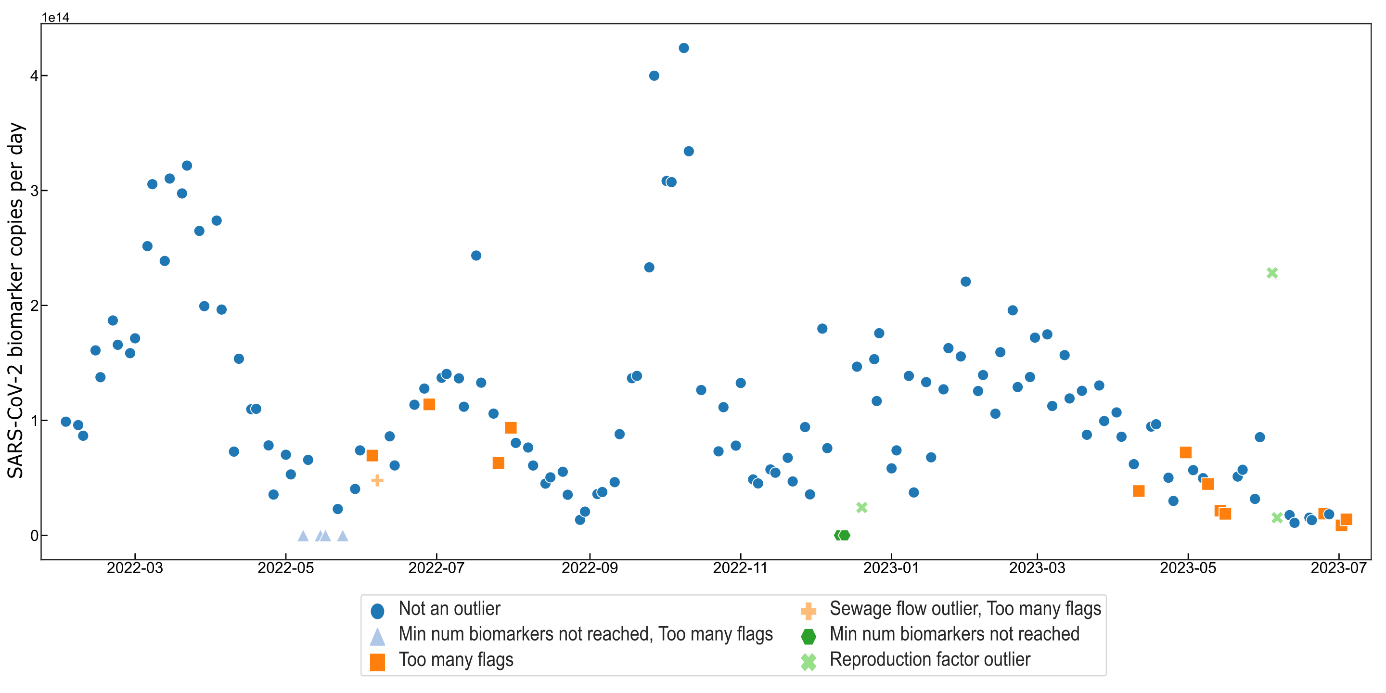
**Supplement S9:** Outlier detection – Nuremberg (Figure)
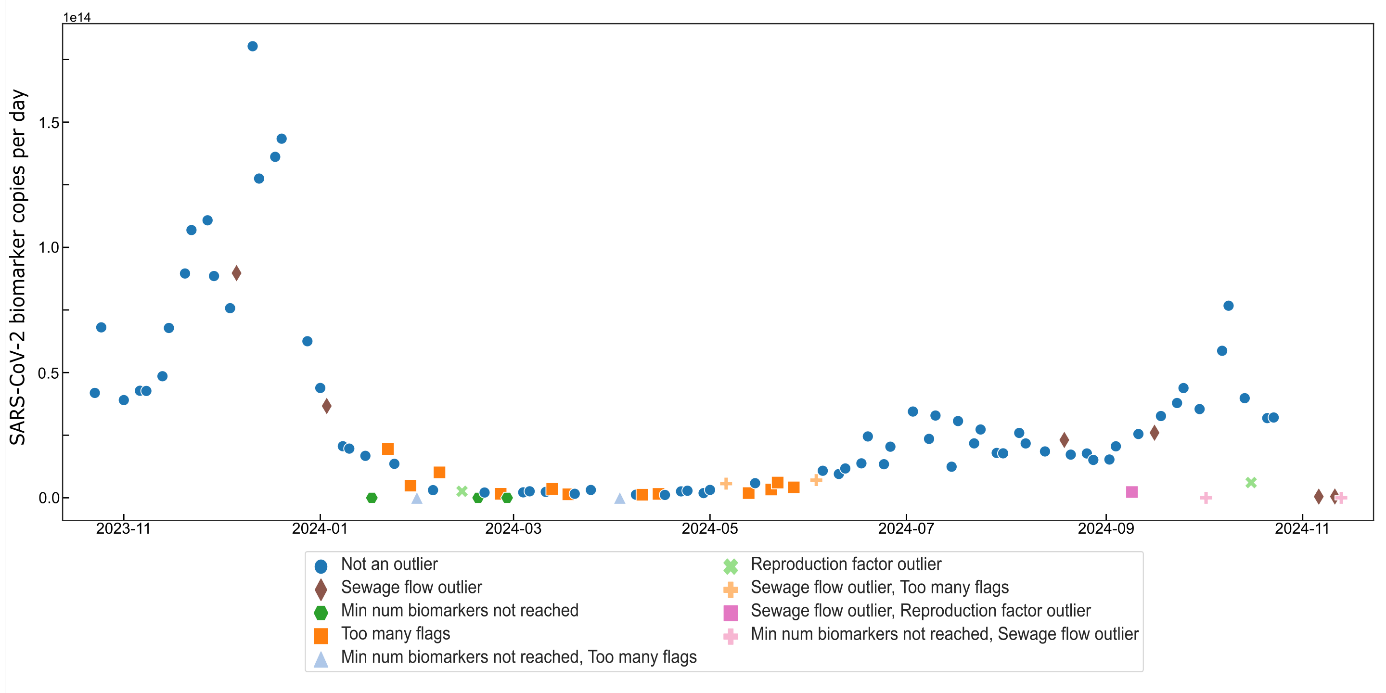

**Supplement S10:** Outlier detection – Karlsruhe (Figure)

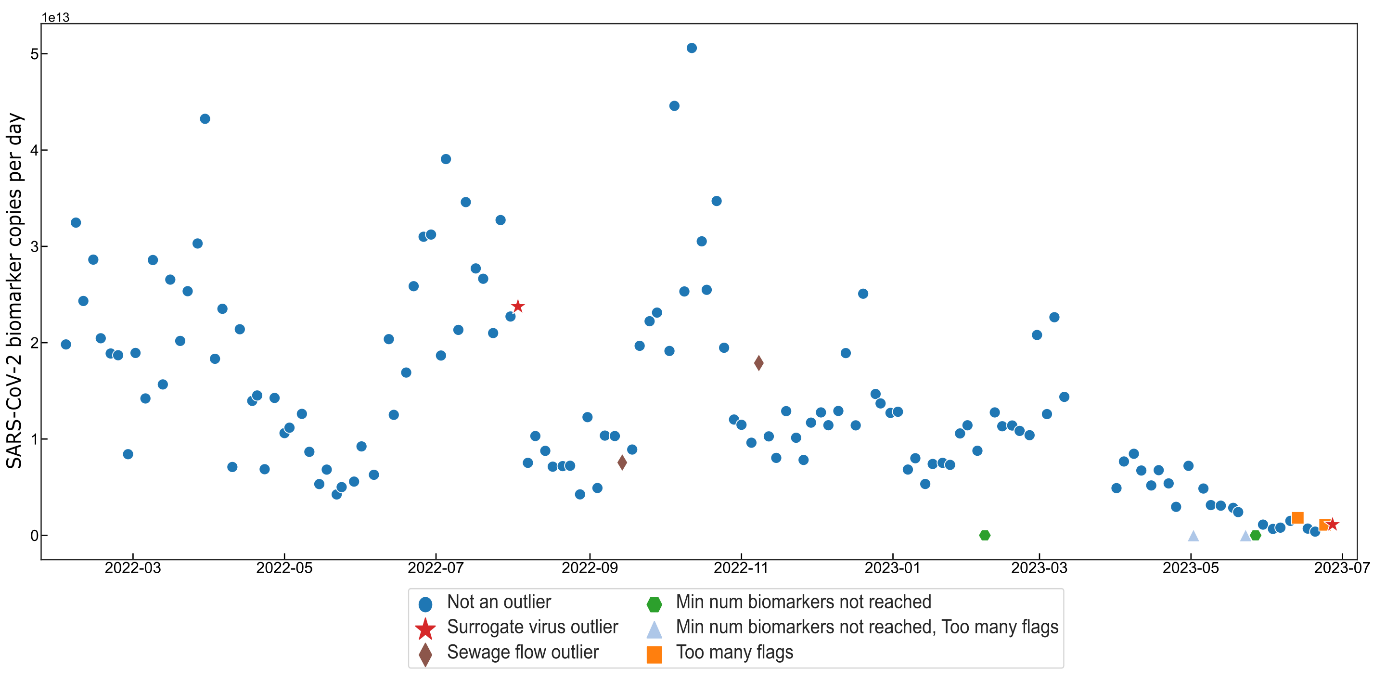

**Supplement S11:** Outlier detection – Königsbrunn (Figure)
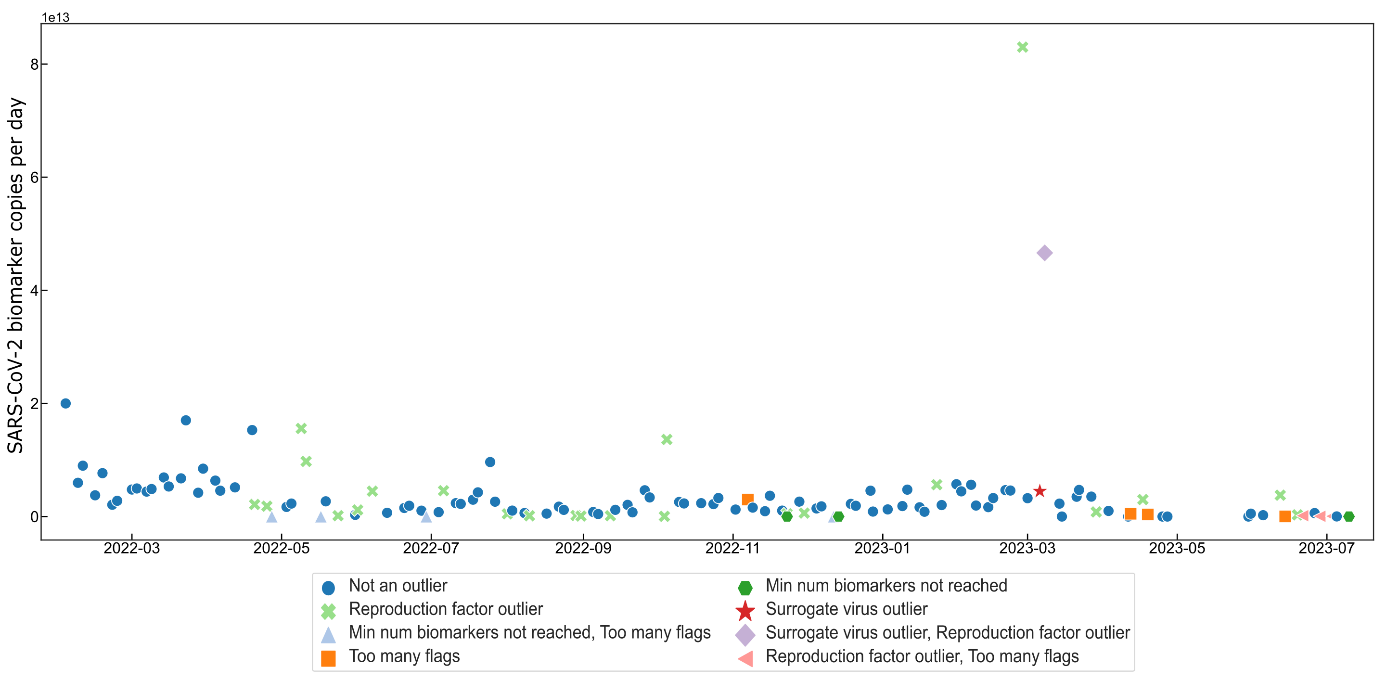

**Supplement S12:** Outlier detection – Berchtesgaden (Figure)
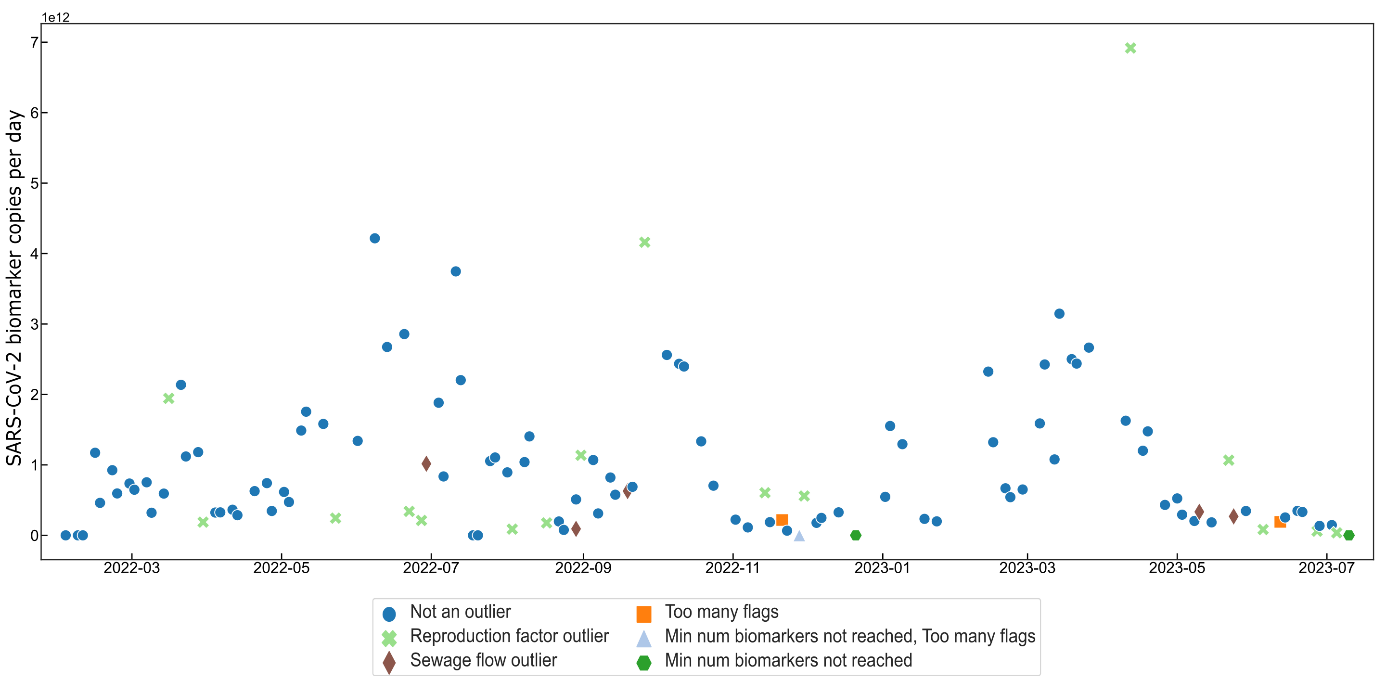

**Supplement S13:** Outlier detection – Ebersberg (Figure)
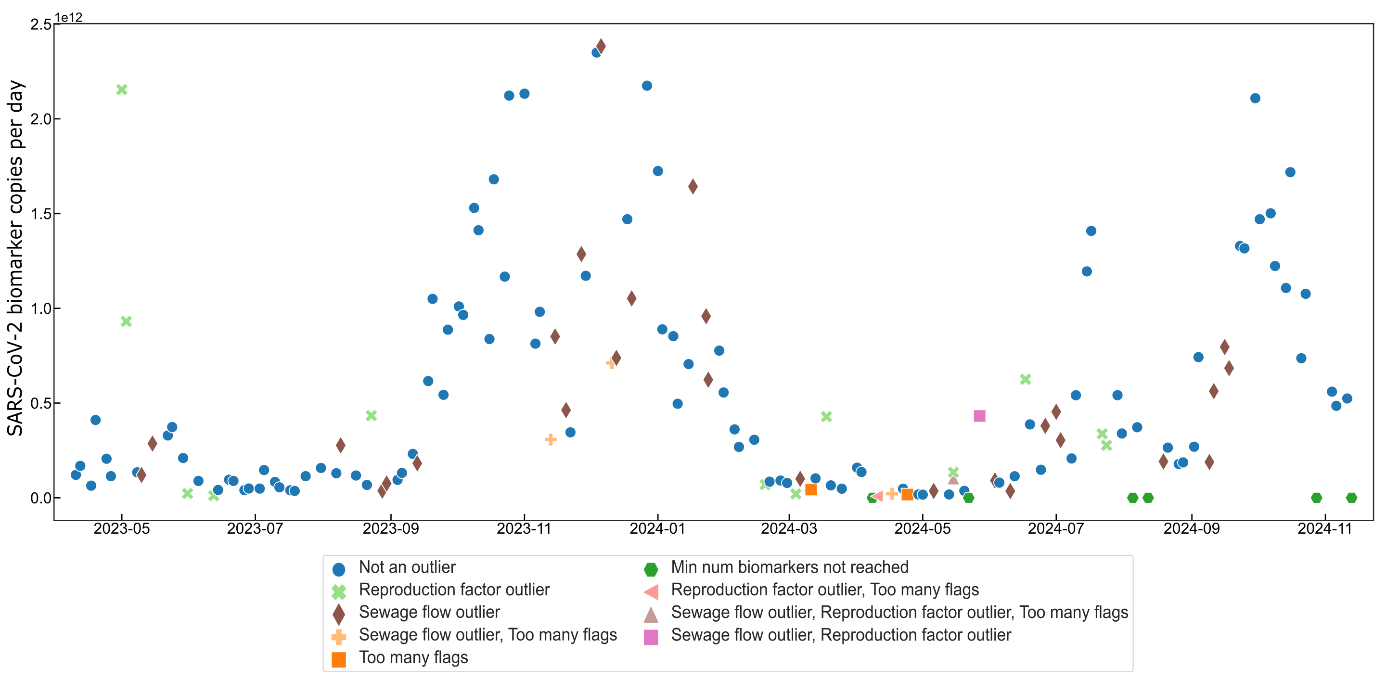

**Supplement S14:** Outlier detection – Piding (Figure)

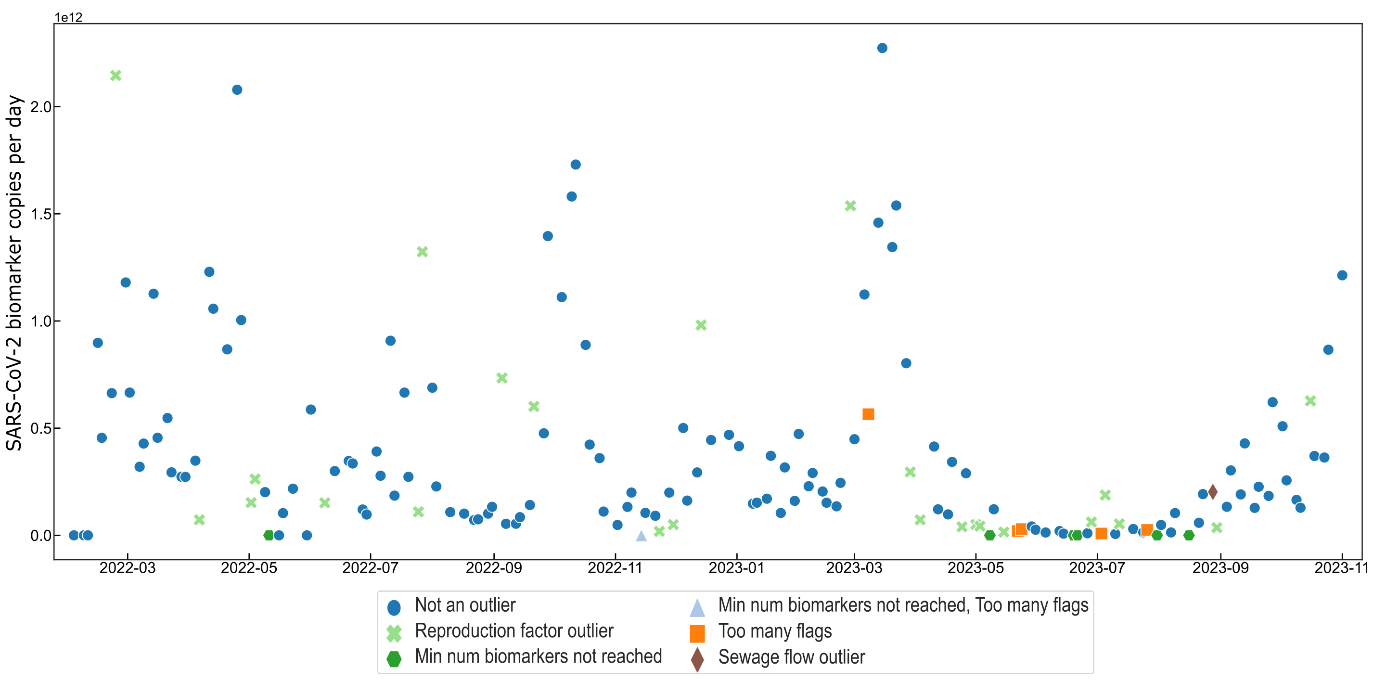

**Supplement S15:** Spearman’s rank-order correlation analysis between wastewater-based epidemiology results and clinical SARS-CoV-2 data for the city of Munich and a Munich city district, tested for various time lags across different SARS-CoV-2 variants and the entire sampling period. The time lag (in days) represents the lead time of wastewater data compared to clinical data, based on the assumption that wastewater data provide earlier results due to rapid sampling, laboratory processing, analysis, and reporting (significance: *** p < 0.001, ** p < 0.01, * p < 0.05). (Table)

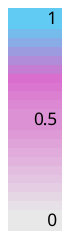

| Biomarker | | Munich | | | | District | | | | Biomarker | | Munich | | | | District | | | |
| --- | --- | --- | --- | --- | --- | --- | --- | --- | --- | --- | --- | --- | --- | --- | --- | --- | --- | --- | --- |
| 7-days  Incidence | | WWTP | | Munich | | District | | Munich | | 7-days  Incidence | | WWTP | | Munich | | District | | Munich | |
| Timelag | |  | | | | | | | | Timelag | |  | | | | | | | |
| Delta | 0 | 0.90 | *** | 0.82 | *** | 0.71 | *** | 0.62 | *** | Omikron BA.1 | 0 | 0.12 |  | 0.05 |  | 0.62 | *** | 0.43 | *** |
|  | 1 | 0.91 | *** | 0.83 | *** | 0.74 | *** | 0.63 | *** |  | 1 | 0.18 |  | 0.08 |  | 0.66 | *** | 0.48 | *** |
|  | 2 | 0.92 | *** | 0.84 | *** | 0.75 | *** | 0.63 | *** |  | 2 | 0.24 | * | 0.12 |  | 0.70 | *** | 0.53 | *** |
|  | 3 | 0.92 | *** | 0.85 | *** | 0.77 | *** | 0.64 | *** |  | 3 | 0.30 | ** | 0.17 |  | 0.74 | *** | 0.57 | *** |
|  | 4 | 0.91 | *** | 0.85 | *** | 0.77 | *** | 0.63 | *** |  | 4 | 0.37 | ** | 0.22 |  | 0.77 | *** | 0.62 | *** |
|  | 5 | 0.89 | *** | 0.84 | *** | 0.77 | *** | 0.63 | *** |  | 5 | 0.45 | *** | 0.27 | * | 0.79 | *** | 0.66 | *** |
|  | 6 | 0.87 | *** | 0.83 | *** | 0.76 | *** | 0.61 | *** |  | 6 | 0.53 | *** | 0.33 | ** | 0.80 | *** | 0.69 | *** |
|  | 7 | 0.84 | *** | 0.83 | *** | 0.74 | *** | 0.60 | *** |  | 7 | 0.60 | *** | 0.39 | *** | 0.80 | *** | 0.72 | *** |
|  | 8 | 0.83 | *** | 0.82 | *** | 0.76 | *** | 0.63 | *** |  | 8 | 0.67 | *** | 0.44 | *** | 0.81 | *** | 0.75 | *** |
|  | 9 | 0.83 | *** | 0.83 | *** | 0.76 | *** | 0.65 | *** |  | 9 | 0.72 | *** | 0.50 | *** | 0.81 | *** | 0.78 | *** |
|  | 10 | 0.83 | *** | 0.83 | *** | 0.76 | *** | 0.66 | *** |  | 10 | 0.76 | *** | 0.57 | *** | 0.81 | *** | 0.80 | *** |
|  | 11 | 0.83 | *** | 0.83 | *** | 0.75 | *** | 0.69 | *** |  | 11 | 0.79 | *** | 0.63 | *** | 0.79 | *** | 0.82 | *** |
|  | 12 | 0.82 | *** | 0.83 | *** | 0.74 | *** | 0.70 | *** |  | 12 | 0.82 | *** | 0.68 | *** | 0.77 | *** | 0.83 | *** |
|  | 13 | 0.81 | *** | 0.83 | *** | 0.72 | *** | 0.71 | *** |  | 13 | 0.83 | *** | 0.73 | *** | 0.75 | *** | 0.83 | *** |
|  | 14 | 0.79 | *** | 0.82 | *** | 0.69 | *** | 0.71 | *** |  | 14 | 0.83 | *** | 0.76 | *** | 0.72 | *** | 0.82 | *** |
|  | 15 | 0.77 | *** | 0.83 | *** | 0.66 | *** | 0.72 | *** |  | 15 | 0.82 | *** | 0.77 | *** | 0.68 | *** | 0.80 | *** |

| Biomarker | | Munich | | | | District | | | | Biomarker | | Munich | | | | District | | | |
| --- | --- | --- | --- | --- | --- | --- | --- | --- | --- | --- | --- | --- | --- | --- | --- | --- | --- | --- | --- |
| 7-days  Incidence | | WWTP | | Munich | | District | | Munich | | 7-days  Incidence | | WWTP | | Munich | | District | | Munich | |
| Timelag | |  | | | | | | | | Timelag | |  | | | | | | | |
| Omikron BA.2 | 0 | 0.96 | *** | 0.91 | *** | 0.93 | *** | 0.91 | *** | Omikron BA.4 and BA.5 | 0 | 0.94 | *** | 0.88 | *** | 0.79 | *** | 0.78 | *** |
|  | 1 | 0.96 | *** | 0.93 | *** | 0.94 | *** | 0.92 | *** |  | 1 | 0.95 | *** | 0.89 | *** | 0.79 | *** | 0.76 | *** |
|  | 2 | 0.96 | *** | 0.94 | *** | 0.95 | *** | 0.93 | *** |  | 2 | 0.94 | *** | 0.90 | *** | 0.78 | *** | 0.75 | *** |
|  | 3 | 0.97 | *** | 0.95 | *** | 0.95 | *** | 0.94 | *** |  | 3 | 0.94 | *** | 0.91 | *** | 0.77 | *** | 0.72 | *** |
|  | 4 | 0.97 | *** | 0.95 | *** | 0.96 | *** | 0.94 | *** |  | 4 | 0.92 | *** | 0.91 | *** | 0.75 | *** | 0.70 | *** |
|  | 5 | 0.97 | *** | 0.96 | *** | 0.96 | *** | 0.95 | *** |  | 5 | 0.91 | *** | 0.91 | *** | 0.73 | *** | 0.68 | *** |
|  | 6 | 0.97 | *** | 0.96 | *** | 0.96 | *** | 0.95 | *** |  | 6 | 0.89 | *** | 0.90 | *** | 0.70 | *** | 0.66 | *** |
|  | 7 | 0.97 | *** | 0.96 | *** | 0.97 | *** | 0.96 | *** |  | 7 | 0.86 | *** | 0.90 | *** | 0.68 | *** | 0.64 | *** |
|  | 8 | 0.97 | *** | 0.97 | *** | 0.97 | *** | 0.96 | *** |  | 8 | 0.83 | *** | 0.89 | *** | 0.64 | *** | 0.60 | *** |
|  | 9 | 0.97 | *** | 0.98 | *** | 0.97 | *** | 0.96 | *** |  | 9 | 0.79 | *** | 0.86 | *** | 0.61 | *** | 0.56 | *** |
|  | 10 | 0.96 | *** | 0.98 | *** | 0.97 | *** | 0.97 | *** |  | 10 | 0.75 | *** | 0.82 | *** | 0.57 | *** | 0.51 | *** |
|  | 11 | 0.96 | *** | 0.98 | *** | 0.96 | *** | 0.97 | *** |  | 11 | 0.70 | *** | 0.78 | *** | 0.53 | *** | 0.46 | *** |
|  | 12 | 0.95 | *** | 0.97 | *** | 0.96 | *** | 0.97 | *** |  | 12 | 0.65 | *** | 0.73 | *** | 0.49 | *** | 0.41 | *** |
|  | 13 | 0.95 | *** | 0.97 | *** | 0.95 | *** | 0.96 | *** |  | 13 | 0.59 | *** | 0.69 | *** | 0.45 | *** | 0.35 | ** |
|  | 14 | 0.94 | *** | 0.96 | *** | 0.94 | *** | 0.96 | *** |  | 14 | 0.53 | *** | 0.64 | *** | 0.40 | *** | 0.28 | * |
|  | 15 | 0.94 | *** | 0.96 | *** | 0.93 | *** | 0.96 | *** |  | 15 | 0.46 | *** | 0.58 | *** | 0.35 | ** | 0.21 |  |

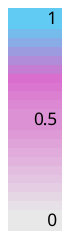

| Biomarker | | Munich | | | | District | | | | Biomarker | | Munich | | | | District | | | |
| --- | --- | --- | --- | --- | --- | --- | --- | --- | --- | --- | --- | --- | --- | --- | --- | --- | --- | --- | --- |
| 7-days  Incidence | | WWTP | | Munich | | District | | Munich | | 7-days  Incidence | | WWTP | | Munich | | District | | Munich | |
| Timelag | |  | | | | | | | | Timelag | |  | | | | | | | |
| Omikron BA.5 and BQ | 0 | 0.71 | *** | 0.69 | *** | 0.80 | *** | 0.88 | *** | Whole period | 0 | 0.81 | *** | 0.78 | *** | 0.79 | *** | 0.77 | *** |
|  | 1 | 0.71 | *** | 0.69 | *** | 0.81 | *** | 0.89 | *** |  | 1 | 0.82 | *** | 0.79 | *** | 0.80 | *** | 0.78 | *** |
|  | 2 | 0.70 | *** | 0.70 | *** | 0.83 | *** | 0.89 | *** |  | 2 | 0.82 | *** | 0.80 | *** | 0.81 | *** | 0.78 | *** |
|  | 3 | 0.68 | *** | 0.70 | *** | 0.84 | *** | 0.88 | *** |  | 3 | 0.82 | *** | 0.80 | *** | 0.82 | *** | 0.79 | *** |
|  | 4 | 0.67 | *** | 0.69 | *** | 0.84 | *** | 0.87 | *** |  | 4 | 0.81 | *** | 0.81 | *** | 0.82 | *** | 0.79 | *** |
|  | 5 | 0.65 | *** | 0.68 | *** | 0.85 | *** | 0.85 | *** |  | 5 | 0.81 | *** | 0.80 | *** | 0.82 | *** | 0.78 | *** |
|  | 6 | 0.63 | *** | 0.66 | *** | 0.85 | *** | 0.83 | *** |  | 6 | 0.80 | *** | 0.80 | *** | 0.82 | *** | 0.78 | *** |
|  | 7 | 0.61 | *** | 0.64 | *** | 0.84 | *** | 0.81 | *** |  | 7 | 0.79 | *** | 0.80 | *** | 0.82 | *** | 0.77 | *** |
|  | 8 | 0.59 | *** | 0.63 | *** | 0.84 | *** | 0.79 | *** |  | 8 | 0.78 | *** | 0.79 | *** | 0.81 | *** | 0.77 | *** |
|  | 9 | 0.57 | *** | 0.61 | *** | 0.84 | *** | 0.77 | *** |  | 9 | 0.77 | *** | 0.79 | *** | 0.79 | *** | 0.75 | *** |
|  | 10 | 0.54 | *** | 0.58 | *** | 0.83 | *** | 0.75 | *** |  | 10 | 0.76 | *** | 0.78 | *** | 0.78 | *** | 0.74 | *** |
|  | 11 | 0.51 | *** | 0.55 | *** | 0.82 | *** | 0.72 | *** |  | 11 | 0.74 | *** | 0.76 | *** | 0.76 | *** | 0.72 | *** |
|  | 12 | 0.47 | *** | 0.52 | *** | 0.80 | *** | 0.68 | *** |  | 12 | 0.72 | *** | 0.75 | *** | 0.73 | *** | 0.70 | *** |
|  | 13 | 0.44 | *** | 0.48 | *** | 0.77 | *** | 0.63 | *** |  | 13 | 0.70 | *** | 0.73 | *** | 0.71 | *** | 0.67 | *** |
|  | 14 | 0.40 | *** | 0.44 | *** | 0.74 | *** | 0.58 | *** |  | 14 | 0.68 | *** | 0.71 | *** | 0.68 | *** | 0.65 | *** |
|  | 15 | 0.35 | *** | 0.40 | *** | 0.70 | *** | 0.52 | *** |  | 15 | 0.65 | *** | 0.69 | *** | 0.64 | *** | 0.62 | *** |

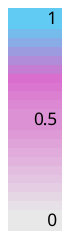

**Supplement S16:** Spearman’s rank-order correlation analysis between wastewater-based epidemiology results and clinical SARS-CoV-2 data for the city of Augsburg and the communities of Königsbrunn and Stadtbergen in the county of Augsburg, tested for various time lags across different SARS-CoV-2 variants and the entire sampling period. The time lag (in days) represents the lead time of wastewater data compared to clinical data, based on the assumption that wastewater data provide earlier results due to rapid sampling, laboratory processing, analysis, and reporting (significance: *** p < 0.001, ** p < 0.01, * p < 0.05).

| SARS-CoV-2 biomarker | | Augsburg | | Königsbrunn | | | | Stadtbergen | | | |
| --- | --- | --- | --- | --- | --- | --- | --- | --- | --- | --- | --- |
| 7-days  Incidence | | Augsburg | | Königsbrunn | | Augsburg | | Stadtbergen | | Augsburg | |
| Timelag | |  | | | | | | | | | |
| Delta | 0 | 0.83 | *** | 0.83 | *** | 0.74 | *** | 0.71 | *** | 0.71 | *** |
|  | 1 | 0.83 | *** | 0.84 | *** | 0.77 | *** | 0.70 | *** | 0.72 | *** |
|  | 2 | 0.83 | *** | 0.83 | *** | 0.79 | *** | 0.70 | *** | 0.72 | *** |
|  | 3 | 0.83 | *** | 0.83 | *** | 0.81 | *** | 0.70 | *** | 0.72 | *** |
|  | 4 | 0.82 | *** | 0.82 | *** | 0.84 | *** | 0.70 | *** | 0.72 | *** |
|  | 5 | 0.82 | *** | 0.83 | *** | 0.86 | *** | 0.68 | *** | 0.73 | *** |
|  | 6 | 0.81 | *** | 0.83 | *** | 0.88 | *** | 0.66 | *** | 0.73 | *** |
|  | 7 | 0.81 | *** | 0.84 | *** | 0.90 | *** | 0.65 | *** | 0.72 | *** |
|  | 8 | 0.80 | *** | 0.84 | *** | 0.91 | *** | 0.64 | *** | 0.71 | *** |
|  | 9 | 0.80 | *** | 0.84 | *** | 0.91 | *** | 0.63 | *** | 0.71 | *** |
|  | 10 | 0.79 | *** | 0.84 | *** | 0.91 | *** | 0.61 | *** | 0.70 | *** |
|  | 11 | 0.78 | *** | 0.82 | *** | 0.90 | *** | 0.60 | *** | 0.68 | *** |
|  | 12 | 0.77 | *** | 0.81 | *** | 0.89 | *** | 0.58 | *** | 0.66 | *** |
|  | 13 | 0.76 | *** | 0.79 | *** | 0.87 | *** | 0.56 | *** | 0.63 | *** |
|  | 14 | 0.75 | *** | 0.76 | *** | 0.86 | *** | 0.55 | *** | 0.61 | *** |
|  | 15 | 0.74 | *** | 0.74 | *** | 0.85 | *** | 0.54 | *** | 0.58 | *** |

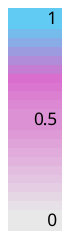

| SARS-CoV-2 biomarker | | Augsburg | | Königsbrunn | | | | Stadtbergen | | | |
| --- | --- | --- | --- | --- | --- | --- | --- | --- | --- | --- | --- |
| 7-days  Incidence | | Augsburg | | Königsbrunn | | Augsburg | | Stadtbergen | | Augsburg | |
| Timelag | |  | | | | | | | | | |
| Omikron BA.1 | 0 | 0.58 | *** | 0.77 | *** | 0.82 | *** | 0.77 | *** | 0.81 | *** |
|  | 1 | 0.61 | *** | 0.81 | *** | 0.85 | *** | 0.75 | *** | 0.79 | *** |
|  | 2 | 0.63 | *** | 0.84 | *** | 0.88 | *** | 0.73 | *** | 0.78 | *** |
|  | 3 | 0.65 | *** | 0.87 | *** | 0.90 | *** | 0.71 | *** | 0.76 | *** |
|  | 4 | 0.66 | *** | 0.90 | *** | 0.91 | *** | 0.68 | *** | 0.74 | *** |
|  | 5 | 0.69 | *** | 0.92 | *** | 0.93 | *** | 0.64 | *** | 0.71 | *** |
|  | 6 | 0.71 | *** | 0.94 | *** | 0.93 | *** | 0.59 | *** | 0.68 | *** |
|  | 7 | 0.72 | *** | 0.95 | *** | 0.92 | *** | 0.53 | *** | 0.65 | *** |
|  | 8 | 0.74 | *** | 0.94 | *** | 0.91 | *** | 0.48 | *** | 0.60 | *** |
|  | 9 | 0.74 | *** | 0.93 | *** | 0.88 | *** | 0.42 | *** | 0.54 | *** |
|  | 10 | 0.73 | *** | 0.90 | *** | 0.83 | *** | 0.36 | ** | 0.50 | *** |
|  | 11 | 0.71 | *** | 0.86 | *** | 0.78 | *** | 0.31 | * | 0.45 | *** |
|  | 12 | 0.69 | *** | 0.82 | *** | 0.73 | *** | 0.25 | * | 0.39 | ** |
|  | 13 | 0.65 | *** | 0.77 | *** | 0.66 | *** | 0.18 |  | 0.34 | ** |
|  | 14 | 0.60 | *** | 0.71 | *** | 0.59 | *** | 0.12 |  | 0.29 | * |
|  | 15 | 0.54 | *** | 0.63 | *** | 0.52 | *** | 0.07 |  | 0.22 |  |

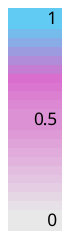

| SARS-CoV-2 biomarker | | Augsburg | | Königsbrunn | | | | Stadtbergen | | | |
| --- | --- | --- | --- | --- | --- | --- | --- | --- | --- | --- | --- |
| 7-days  Incidence | | Augsburg | | Königsbrunn | | Augsburg | | Stadtbergen | | Augsburg | |
| Timelag | |  | | | | | | | | | |
| Omikron BA.2 | 0 | 0.93 | *** | 0.41 | *** | 0.37 | *** | 0.82 | *** | 0.86 | *** |
|  | 1 | 0.94 | *** | 0.39 | *** | 0.34 | ** | 0.80 | *** | 0.85 | *** |
|  | 2 | 0.95 | *** | 0.38 | *** | 0.32 | ** | 0.77 | *** | 0.83 | *** |
|  | 3 | 0.96 | *** | 0.36 | *** | 0.29 | ** | 0.76 | *** | 0.82 | *** |
|  | 4 | 0.97 | *** | 0.35 | ** | 0.27 | * | 0.74 | *** | 0.80 | *** |
|  | 5 | 0.97 | *** | 0.33 | ** | 0.25 | * | 0.71 | *** | 0.79 | *** |
|  | 6 | 0.97 | *** | 0.30 | ** | 0.24 | * | 0.70 | *** | 0.77 | *** |
|  | 7 | 0.96 | *** | 0.28 | * | 0.22 | * | 0.69 | *** | 0.76 | *** |
|  | 8 | 0.95 | *** | 0.23 | * | 0.17 |  | 0.65 | *** | 0.74 | *** |
|  | 9 | 0.94 | *** | 0.18 |  | 0.14 |  | 0.63 | *** | 0.71 | *** |
|  | 10 | 0.93 | *** | 0.13 |  | 0.10 |  | 0.60 | *** | 0.69 | *** |
|  | 11 | 0.91 | *** | 0.07 |  | 0.07 |  | 0.57 | *** | 0.67 | *** |
|  | 12 | 0.90 | *** | 0.03 |  | 0.03 |  | 0.55 | *** | 0.64 | *** |
|  | 13 | 0.88 | *** | 0.00 |  | -0.01 |  | 0.52 | *** | 0.63 | *** |
|  | 14 | 0.87 | *** | -0.05 |  | -0.03 |  | 0.50 | *** | 0.62 | *** |
|  | 15 | 0.86 | *** | -0.09 |  | -0.07 |  | 0.49 | *** | 0.60 | *** |

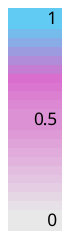

| SARS-CoV-2 biomarker | | Augsburg | | Königsbrunn | | | | Stadtbergen | | | |
| --- | --- | --- | --- | --- | --- | --- | --- | --- | --- | --- | --- |
| 7-days  Incidence | | Augsburg | | Königsbrunn | | Augsburg | | Stadtbergen | | Augsburg | |
| Timelag | |  | | | | | | | | | |
| Omikron BA.4 and BA.5 | 0 | 0.95 | *** | 0.75 | *** | 0.83 | *** | 0.52 | *** | 0.67 | *** |
|  | 1 | 0.97 | *** | 0.75 | *** | 0.86 | *** | 0.56 | *** | 0.70 | *** |
|  | 2 | 0.98 | *** | 0.75 | *** | 0.86 | *** | 0.60 | *** | 0.71 | *** |
|  | 3 | 0.98 | *** | 0.75 | *** | 0.86 | *** | 0.64 | *** | 0.74 | *** |
|  | 4 | 0.97 | *** | 0.73 | *** | 0.86 | *** | 0.67 | *** | 0.76 | *** |
|  | 5 | 0.96 | *** | 0.71 | *** | 0.85 | *** | 0.71 | *** | 0.77 | *** |
|  | 6 | 0.95 | *** | 0.66 | *** | 0.83 | *** | 0.75 | *** | 0.78 | *** |
|  | 7 | 0.94 | *** | 0.64 | *** | 0.82 | *** | 0.78 | *** | 0.79 | *** |
|  | 8 | 0.91 | *** | 0.62 | *** | 0.79 | *** | 0.79 | *** | 0.79 | *** |
|  | 9 | 0.88 | *** | 0.59 | *** | 0.76 | *** | 0.80 | *** | 0.78 | *** |
|  | 10 | 0.85 | *** | 0.57 | *** | 0.73 | *** | 0.81 | *** | 0.78 | *** |
|  | 11 | 0.81 | *** | 0.54 | *** | 0.69 | *** | 0.81 | *** | 0.77 | *** |
|  | 12 | 0.76 | *** | 0.51 | *** | 0.64 | *** | 0.80 | *** | 0.77 | *** |
|  | 13 | 0.70 | *** | 0.44 | *** | 0.59 | *** | 0.79 | *** | 0.76 | *** |
|  | 14 | 0.65 | *** | 0.34 | ** | 0.52 | *** | 0.77 | *** | 0.75 | *** |
|  | 15 | 0.58 | *** | 0.27 | * | 0.44 | *** | 0.75 | *** | 0.73 | *** |

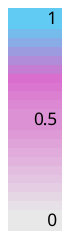

| SARS-CoV-2 biomarker | | Augsburg | | Königsbrunn | | | | Stadtbergen | | | |
| --- | --- | --- | --- | --- | --- | --- | --- | --- | --- | --- | --- |
| 7-days  Incidence | | Augsburg | | Königsbrunn | | Augsburg | | Stadtbergen | | Augsburg | |
| Timelag | |  | | | | | | | | | |
| Omikron BA.5 and BQ | 0 | 0.73 | *** | 0.79 | *** | 0.81 | *** | 0.83 | *** | 0.82 | *** |
|  | 1 | 0.74 | *** | 0.80 | *** | 0.81 | *** | 0.82 | *** | 0.82 | *** |
|  | 2 | 0.73 | *** | 0.81 | *** | 0.81 | *** | 0.80 | *** | 0.81 | *** |
|  | 3 | 0.73 | *** | 0.80 | *** | 0.80 | *** | 0.78 | *** | 0.80 | *** |
|  | 4 | 0.71 | *** | 0.80 | *** | 0.78 | *** | 0.77 | *** | 0.79 | *** |
|  | 5 | 0.70 | *** | 0.79 | *** | 0.76 | *** | 0.75 | *** | 0.77 | *** |
|  | 6 | 0.68 | *** | 0.77 | *** | 0.75 | *** | 0.71 | *** | 0.75 | *** |
|  | 7 | 0.66 | *** | 0.74 | *** | 0.73 | *** | 0.67 | *** | 0.73 | *** |
|  | 8 | 0.63 | *** | 0.70 | *** | 0.70 | *** | 0.62 | *** | 0.69 | *** |
|  | 9 | 0.60 | *** | 0.66 | *** | 0.67 | *** | 0.57 | *** | 0.66 | *** |
|  | 10 | 0.57 | *** | 0.62 | *** | 0.62 | *** | 0.52 | *** | 0.62 | *** |
|  | 11 | 0.53 | *** | 0.57 | *** | 0.57 | *** | 0.47 | *** | 0.58 | *** |
|  | 12 | 0.49 | *** | 0.52 | *** | 0.52 | *** | 0.42 | *** | 0.54 | *** |
|  | 13 | 0.44 | *** | 0.47 | *** | 0.46 | *** | 0.37 | *** | 0.50 | *** |
|  | 14 | 0.39 | *** | 0.42 | *** | 0.40 | *** | 0.34 | *** | 0.45 | *** |
|  | 15 | 0.34 | *** | 0.36 | *** | 0.34 | *** | 0.30 | ** | 0.41 | *** |

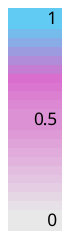

| SARS-CoV-2 biomarker | | Augsburg | | Königsbrunn | | | | Stadtbergen | | | |
| --- | --- | --- | --- | --- | --- | --- | --- | --- | --- | --- | --- |
| 7-days  Incidence | | Augsburg | | Königsbrunn | | Augsburg | | Stadtbergen | | Augsburg | |
| Timelag | |  | | | | | | | | | |
| low incidence (from Dec 2022) | 0 | -0.10 |  | -0.41 | *** | -0.47 | *** | -0.15 |  | -0.30 | ** |
|  | 1 | -0.08 |  | -0.42 | *** | -0.44 | *** | -0.13 |  | -0.25 | * |
|  | 2 | -0.06 |  | -0.45 | *** | -0.42 | *** | -0.08 |  | -0.21 | * |
|  | 3 | -0.03 |  | -0.46 | *** | -0.39 | *** | -0.06 |  | -0.17 |  |
|  | 4 | -0.02 |  | -0.50 | *** | -0.37 | *** | -0.03 |  | -0.14 |  |
|  | 5 | -0.01 |  | -0.49 | *** | -0.34 | ** | -0.02 |  | -0.11 |  |
|  | 6 | 0.00 |  | -0.50 | *** | -0.34 | ** | 0.00 |  | -0.09 |  |
|  | 7 | 0.01 |  | -0.50 | *** | -0.32 | ** | 0.02 |  | -0.08 |  |
|  | 8 | 0.03 |  | -0.50 | *** | -0.30 | ** | 0.01 |  | -0.10 |  |
|  | 9 | 0.05 |  | -0.48 | *** | -0.30 | ** | -0.03 |  | -0.08 |  |
|  | 10 | 0.06 |  | -0.46 | *** | -0.29 | ** | -0.08 |  | -0.07 |  |
|  | 11 | 0.07 |  | -0.41 | *** | -0.28 | * | -0.13 |  | -0.05 |  |
|  | 12 | 0.05 |  | -0.36 | *** | -0.27 | * | -0.21 |  | -0.02 |  |
|  | 13 | 0.05 |  | -0.33 | ** | -0.23 | * | -0.33 | ** | 0.02 |  |
|  | 14 | 0.04 |  | -0.32 | ** | -0.20 |  | -0.43 | *** | 0.06 |  |
|  | 15 | 0.03 |  | -0.31 | ** | -0.14 |  | -0.50 | *** | 0.10 |  |

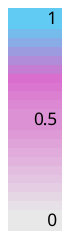

| SARS-CoV-2 biomarker | | Augsburg | | Königsbrunn | | | | Stadtbergen | | | |
| --- | --- | --- | --- | --- | --- | --- | --- | --- | --- | --- | --- |
| 7-days  Incidence | | Augsburg | | Königsbrunn | | Augsburg | | Stadtbergen | | Augsburg | |
| Timelag | |  | | | | | | | | | |
| Whole period | 0 | 0.52 | *** | 0.61 | *** | 0.55 | *** | 0.59 | *** | 0.50 | *** |
|  | 1 | 0.52 | *** | 0.60 | *** | 0.55 | *** | 0.58 | *** | 0.50 | *** |
|  | 2 | 0.52 | *** | 0.59 | *** | 0.54 | *** | 0.57 | *** | 0.50 | *** |
|  | 3 | 0.51 | *** | 0.59 | *** | 0.53 | *** | 0.57 | *** | 0.49 | *** |
|  | 4 | 0.51 | *** | 0.58 | *** | 0.52 | *** | 0.56 | *** | 0.49 | *** |
|  | 5 | 0.50 | *** | 0.56 | *** | 0.51 | *** | 0.56 | *** | 0.48 | *** |
|  | 6 | 0.50 | *** | 0.55 | *** | 0.50 | *** | 0.55 | *** | 0.47 | *** |
|  | 7 | 0.49 | *** | 0.53 | *** | 0.48 | *** | 0.54 | *** | 0.46 | *** |
|  | 8 | 0.48 | *** | 0.52 | *** | 0.47 | *** | 0.52 | *** | 0.45 | *** |
|  | 9 | 0.48 | *** | 0.51 | *** | 0.46 | *** | 0.51 | *** | 0.44 | *** |
|  | 10 | 0.47 | *** | 0.49 | *** | 0.44 | *** | 0.50 | *** | 0.43 | *** |
|  | 11 | 0.46 | *** | 0.47 | *** | 0.43 | *** | 0.48 | *** | 0.42 | *** |
|  | 12 | 0.45 | *** | 0.46 | *** | 0.41 | *** | 0.47 | *** | 0.41 | *** |
|  | 13 | 0.44 | *** | 0.44 | *** | 0.39 | *** | 0.45 | *** | 0.40 | *** |
|  | 14 | 0.43 | *** | 0.42 | *** | 0.37 | *** | 0.44 | *** | 0.39 | *** |
|  | 15 | 0.41 | *** | 0.40 | *** | 0.36 | *** | 0.42 | *** | 0.38 | *** |

**Supplement S17: Frequently Asked Questions (FAQ): How to use and interpret the ANNA-WES dashboard**

When are the dashboards updated?

The ANNA-WES dashboard is updated as soon as possible after sampling. The wastewater samples are taken on Monday and Wednesday. A time delay occurs due to sample transport to the laboratory and the analysis in the laboratory. In the fastest way, the biomarker findings can be reported one day after sampling; Generally, the dashboards are updated three to four days after sampling. If errors occur during sampling or analysis in the laboratory, the reporting can also be omitted. The dashboard on Covid-19 cases is updated daily (Mon-Fri), depending on available capacity.

What is a biomarker and which biomarker is used for wastewater monitoring of SARS-CoV-2?

A biomarker is a biological characteristic that can be measured and evaluated. Body temperature, for example, is a biomarker for fever. In wastewater monitoring of SARS-CoV-2, several genes of the virus are used as biomarkers. Infected people excrete the viruses in their stools. Although the viruses are no longer infectious, they are still detectable.

What is wastewater?

Wastewater is water that has been altered by domestic, commercial, agricultural, or other uses. Wastewater is collected in the sewage system and transported to the treatment plant via sewers. There it is purified and discharged back into the water.

Why is the wastewater sampled?

Studies have confirmed that people infected with SARS-CoV-2 excrete the virus in their stool. Everyone needs to go to the toilet, so wastewater monitoring also detects people who are asymptomatic or who are not registered due to a lack of testing and reporting capacity. Finally, wastewater monitoring is not dependent on test rates. The values measured in wastewater objectively reflect the incidence of infection and should be seen as additional information.

What is a wastewater catchment area?

The wastewater catchment area is the area in which all houses connected to the sewers drain to the same wastewater treatment plant.

Where and how are the wastewater samples taken?

The wastewater samples for SARS-CoV-2 wastewater monitoring are taken at the wastewater treatment plant or, in exceptional cases, directly in the sewer. Samples are usually taken over 24 hours. Samples are taken at regular intervals and mixed. In this way, a representative cross-section of the entire day is obtained. At the end of the 24 hours, the mixed sample is analyzed in the laboratory for the SARS-CoV-2 virus.

How are the wastewater samples analyzed in the laboratory?

The wastewater samples are analyzed in the laboratory using a PCR analysis, similar to the common PCR tests for testing people. Before the actual PCR analysis, the wastewater sample is prepared to extract the RNA biomarkers from the wastewater sample. This serves to prepare the sample for the PCR analysis, so to speak. In the PCR, the individual gene fragments of SARS-CoV-2 are then detected and quantified. For the analysis, the average of several genes is calculated as gene copies per ml of wastewater.

What does the unit gene copies per ml (gene copies/ml) mean?

The unit of the biomarker findings of SARS-CoV-2 is given in gene copies/ml in the wastewater dashboard. The unit gene copies/ml is the unit of PCR analysis and is explained by the PCR analysis procedure. In a PCR analysis, RNA strands are duplicated and the gene copies originally present in the wastewater are then quantified.

What are the uncertainties in wastewater monitoring of SARS-CoV-2?

Wastewater monitoring of SARS-CoV-2 is a new field of research. It has not been conclusively researched which factors affect biomarker concentrations and how. Known uncertainties arise, for example, from the dwell time of the wastewater in the sewer network, i.e. how long the wastewater flows in the sewer system. The SARS-CoV-2 virus fragments contained in the wastewater partially degrade in the wastewater. However, the degradation rate in relation to the retention time in the sewer system is not known. The dilution of wastewater due to precipitation or melting snow also affects the biomarker concentrations. Further research is needed, for example, into the excretion behavior of viruses from an infected person during the course of the infection. It is currently not known at what point during the infection how many viruses are excreted. Despite these uncertainties, research has now progressed to such an extent that the analysis of SARS-CoV-2 biomarkers in wastewater, in addition to clinical testing, can be used to estimate the occurrence of infections.

The biomarker findings are displayed in the dashboard as individual findings and as a moving average. What is the difference?

An individual result is determined from the average of the gene copies per gene reported by the laboratory on the sampling day, with subsequent normalization and considering external influences such as precipitation. Normalization is used here to correct the measured values concerning dilution due to rainwater, snowmelt, penetrating groundwater, etc. For this purpose, the measured SARS-CoV-2 biomarkers are offset against the values of a normalization parameter, thus "normalized". This normalization parameter can be the volume of water flowing into the wastewater treatment plant or a chemical or biological parameter that indicates the faecal load in the wastewater.

The moving average is in turn formed from three consecutive individual findings. "Moving" means that the window used for the calculation shifts. The window containing the values of the individual findings to be considered is shifted with an overlap. This means that the last value is repeatedly deleted from the window under consideration and the first value after the window is added. These values are then used to calculate a new mean value. By using the moving average, strong fluctuations in the individual findings are smoothed out and trends become more visible.

Which graphs in the wastewater dashboard refer to individual biomarker findings and which refer to the moving average?

The tile with the + and - signs in the dashboard shows the change in the moving average of the SARS-CoV-2 biomarker results. A constant level "+/-" is defined as a percentage change of less than 19 % from the last to the previous mean value of the findings. If the change is greater than 19 %, an increased level is symbolized with a "+" or, correspondingly, a decreased level with a "-". The moving average might fall even though an individual finding has risen compared to the previous individual finding.

The bar chart "details" shows the moving averages on the respective sampling day.

In the "timeline" diagram, the individual findings are shown as points and the moving average is shown as a line.

What is the explanation for the "Change" tile with the plus and minus sign in the wastewater dashboard?

The signs symbolize the change in biomarker findings according to the selected measurement area.

A constant level "+/-" is defined as a percentage change of less than 19% from the last to the previous mean value of the findings. If the change is greater, an increased level "+" or correspondingly a decreased level "-" is displayed.

What is the explanation for the "Details" tile with the bar charts in the wastewater dashboard?

The moving averages of the SARS-CoV-2 biomarker findings over the last 30 days are shown. In addition to the display of the change, the extent to which the findings rise or fall in relation to each other is also shown.

The moving average is calculated from five consecutive findings. A distinction is made as to whether the values are considered reliable or not. In the latter case, they are visible in the "Outliers" tab at the bottom of the diagram. In addition, all measured values are initially classified as unknown concerning qualification. These values are displayed via the Details tab.

The mean values of the biomarkers are shown as load or concentration depending on the normalization method. The unit of measurement for the load is gene copies/d. This unit of measurement applies to the normalization method "Effluent". For individual measuring points (e.g. from the Berchtesgadener Land region), normalization is carried out using the "BGL" method. In this case, the values are given in the unit of the PCR measurement as gene copies/ml, even if the value axis shows a different unit.

What is the explanation for the "timeline" diagram with the individual findings and the moving average in the wastewater dashboard?

The temporal course of the SARS-CoV-2 biomarker in wastewater at the selected measuring point is shown. The infection history of the associated wastewater catchment area is subsumed at the measuring point.

The mean values of the biomarkers are shown as a load or concentration depending on the normalization method. Biomarkers are specific target genes for the clear identification of SARS-CoV-2. The unit of measurement for the load is gene copies/d. This unit of measurement applies to the normalization method "effluent". For individual measuring points (e.g. from the Berchtesgadener Land region), normalization is carried out using the "BGL" method. In this case, the values are given in the unit of the PCR measurement as gene copies/ml, even if the value axis shows a different unit.

The individual findings on a sampling day are shown as dots in the diagram. The line represents the moving average, which is formed from three consecutive individual results.

Is there a correlation between the biomarker concentration in wastewater and the reported case numbers?

The correlation between the SARS-CoV-2 biomarker concentration in wastewater and COVID-incidences is the subject of research. In the district of Berchtesgadener Land, it should be noted that the case numbers are reported at the municipal or district level, but the wastewater catchment areas usually deviate from these political boundaries. This means that wastewater monitoring does not cover the same individuals as the incidence reports. Consequently, there should be no direct comparison of incidences and wastewater findings.

Rather than concrete incidence figures, the biomarker findings reflect the occurrence of infections in the recorded catchment area. The wastewater findings thus serve as additional information to the clinical tests. Individuals who show asymptomatic courses or who are not registered due to a lack of testing and reporting capacities can also be recorded via wastewater monitoring.

What does the future of SARS-CoV-2 wastewater monitoring look like?

Wastewater monitoring objectively records the entire population, regardless of the current testing rate of the population. It is therefore an instrument for assessing the occurrence of infection with SARS-CoV-2. In addition, wastewater monitoring via biomarkers can also be a possible instrument for assessing public health in the case of other viruses or, for example, the spread of antibiotic resistance.

The FAQs are based on the Q&A on SARS-CoV-2 wastewater monitoring published by the WHO in 2022 and the National Institute for Public Health and the Environment (RIVM).
